# Lifetime depression and age-related changes in body composition, cardiovascular function, grip strength and lung function: sex-specific analyses in the UK Biobank

**DOI:** 10.1101/2021.02.03.21251076

**Authors:** Julian Mutz, Cathryn M. Lewis

## Abstract

Individuals with depression, on average, die prematurely, have high levels of physical comorbidities and may experience accelerated biological ageing. A greater understanding of age-related changes in physiology could provide novel biological insights that may help inform strategies to mitigate excess mortality in depression. We used generalised additive models to examine age-related changes in 15 cardiovascular, body composition, grip strength and lung function measures, comparing males and females with a lifetime history of depression to healthy controls. The main dataset included 342,393 adults (mean age = 55.87 years, SD = 8.09; 52.61% females). We found statistically significant case-control differences for most physiological measures. There was some evidence that age-related changes in body composition, cardiovascular function, lung function and heel bone mineral density followed different trajectories in depression. These differences did not uniformly narrow or widen with age and differed by sex. For example, BMI in female cases was 1.1 kg/m^2^ higher at age 40 and this difference narrowed to 0.4 kg/m^2^ at age 70. In males, systolic blood pressure was 1 mmHg lower in depression cases at age 45 and this difference widened to 2.5 mmHg at age 65. These findings suggest that targeted screening for physiological function in middle-aged and older adults with depression is warranted to potentially mitigate excess mortality.

## Introduction

Individuals with mental disorders, on average, die prematurely [1] and have high levels of physical comorbidities [2]. Although it is almost universally accepted that health deteriorates as part of the normal ageing process, there is substantial variation in ageing trajectories [3]. Up to a third of older adults experience depression and in many cases show greater levels of functional decline and cognitive impairment [4].

To better understand the heterogeneity in ageing trajectories, it would be useful to objectively assess whether groups or individuals deviate from ‘normal’ age-related changes. To date, no single biological marker that fully captures the ageing process has been discovered, although several candidate measures such as telomere attrition have been described [5]. The measurement of blood-based biomarkers such as leukocyte telomere length is minimally invasive, relatively costly and time-consuming, especially at population scale. Magnetic resonance imaging, for instance used to derive estimates of brain age [6], is costly and less accessible. Physiological measures such as hand-grip strength can be easily obtained, provide reliable information about age-related functional decline and predict morbidity and mortality risk [7-9].

Several studies have examined age-related variation in these measures [10-12]. However, less is known about the ageing trajectories of individuals with depression and whether similar patterns are observed across different physiological measures. A deeper understanding of age-related changes in physiology in individuals with depression might provide insights into the premature mortality observed in mental disorders and could point towards potential targets for prevention or intervention of accelerated ageing [13].

The aim of this study was to examine associations between age and multiple physiological measures, and to test for differences in these measures between middle-aged and older adults with a lifetime history of depression and healthy controls across age. Given major sex differences in physiology, age-related diseases and life expectancy [14], and in prevalence of depression [15], analyses were conducted separately in males and females. In a separate study, we have examined age-related changes in physiology in bipolar disorder [16].

Using data from up to ∼342,000 UK Biobank participants aged 37-73, the following research questions were examined for each physiological measure assessed during the baseline assessment: Are there average differences between males and females with lifetime depression and healthy controls? Do age-related changes differ between males and females with lifetime depression and healthy controls?

## Results

### Study population

Of the 502,521 UK Biobank participants, 392,467 (78.1%) individuals had complete data on all covariates. We yielded an analytical sample of 342,393 adults after excluding individuals who did not meet our inclusion criteria or whose lifetime depression status could not be determined. Sample sizes for physiological measures that were available in subsets of the UK Biobank population ranged from 120,843 to 224,805 participants (Figure 1).

**Figure 1.**
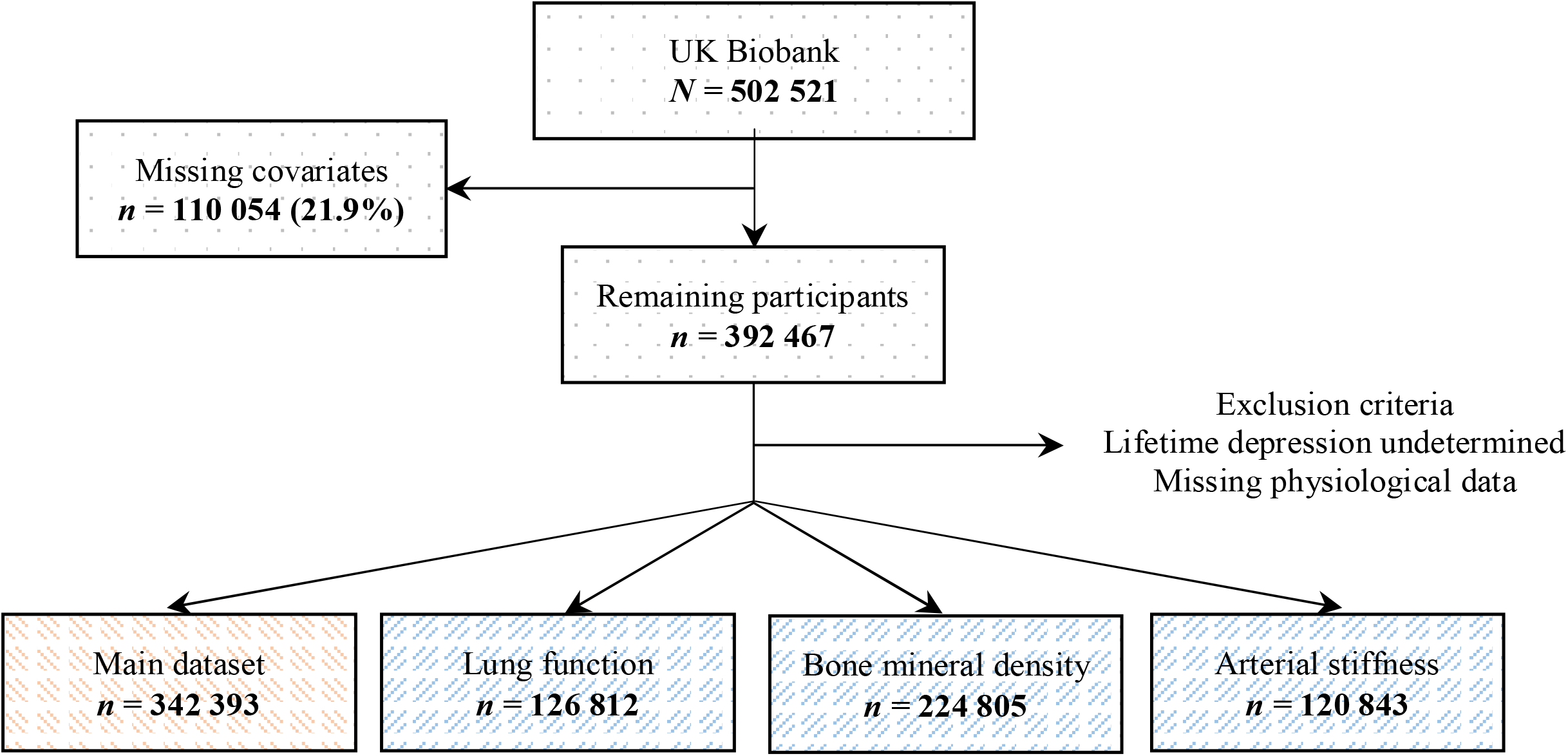
Flowchart of study population. The main dataset included hand-grip strength, blood pressure, pulse rate and measures of body composition.

### Sample characteristics

The average age of participants in our main dataset was 55.87 years (SD = 8.09) and 52.61% of participants were female. About 22.25% of participants (*n* = 87,339) met criteria for lifetime depression, 64.43% (*n* = 56,276) of whom were female. Descriptive statistics and density plots of the physiological measures stratified by sex and lifetime depression status are presented in Table 1 and Supplement 5, respectively. Descriptive statistics of the covariates are presented in Supplement 6.

**Table 1.** Physiological measures

|  |  | Female |  | Male |  |
| --- | --- | --- | --- | --- | --- |
|  | Overall<br>(N=502521) | Healthy control<br>(N=123842) | Depression<br>(N=56276) | Healthy control<br>(N=131212) | Depression<br>(N=31063) |
| **Hand-grip strength** |  |  |  |  |  |
| Mean (SD) | 31.77 (11.34) | 25.10 (6.46) | 24.73 (6.67) | 41.34 (9.09) | 40.37 (9.37) |
| Missing | 3203 |  |  |  |  |
| **Systolic blood pressure** |  |  |  |  |  |
| Mean (SD) | 137.87 (18.65) | 135.07 (19.20) | 132.32 (18.28) | 141.11 (17.38) | 139.02 (16.78) |
| Missing | 1325 |  |  |  |  |
| **Diastolic blood pressure** |  |  |  |  |  |
| Mean (SD) | 82.26 (10.15) | 80.63 (9.98) | 80.26 (9.93) | 84.19 (9.96) | 83.78 (10.02) |
| Missing | 1323 |  |  |  |  |
| **Pulse rate** |  |  |  |  |  |
| Mean (SD) | 69.42 (11.26) | 69.83 (10.38) | 70.05 (10.52) | 67.74 (11.59) | 68.70 (11.99) |
| Missing | 1323 |  |  |  |  |
| **Body mass index** |  |  |  |  |  |
| Mean (SD) | 27.43 (4.80) | 26.63 (4.86) | 27.35 (5.39) | 27.62 (4.01) | 28.08 (4.45) |
| Missing | 3105 |  |  |  |  |
| **Body fat percentage** |  |  |  |  |  |
| Mean (SD) | 31.45 (8.55) | 35.98 (6.77) | 36.87 (7.02) | 24.93 (5.63) | 25.55 (5.88) |
| Missing | 10408 |  |  |  |  |
| **Fat mass** |  |  |  |  |  |
| Mean (SD) | 24.86 (9.57) | 26.12 (9.55) | 27.67 (10.60) | 21.85 (7.87) | 22.88 (8.71) |
| Missing | 10973 |  |  |  |  |
| **Fat-free mass** |  |  |  |  |  |
| Mean (SD) | 53.22 (11.50) | 44.43 (4.85) | 45.05 (5.17) | 63.74 (7.61) | 64.25 (7.93) |
| Missing | 10176 |  |  |  |  |
| **Waist-hip ratio** |  |  |  |  |  |
| Mean (SD) | 0.87 (0.09) | 0.81 (0.07) | 0.82 (0.07) | 0.93 (0.06) | 0.94 (0.07) |
| Missing | 2265 |  |  |  |  |
|  |  | N=43133 | N=19146 | N=52359 | N=12177 |
| **Peak expiratory flow** |  |  |  |  |  |
| Mean (SD) | 407.82 (130.78) | 355.08 (76.97) | 359.35 (76.98) | 511.67 (115.38) | 509.05 (117.30) |
| Missing | 168566 |  |  |  |  |
| **Forced expiratory volume 1s** |  |  |  |  |  |
| Mean (SD) | 2.85 (0.78) | 2.50 (0.53) | 2.53 (0.53) | 3.45 (0.73) | 3.43 (0.75) |
| Missing | 149197 |  |  |  |  |
| **Forced vital capacity** |  |  |  |  |  |
| Mean (SD) | 3.78 (0.98) | 3.28 (0.64) | 3.31 (0.64) | 4.59 (0.88) | 4.57 (0.90) |
| Missing | 149197 |  |  |  |  |
| **FEV<sub>1</sub> / FVC** |  |  |  |  |  |
| Mean (SD) | 0.75 (0.07) | 0.76 (0.06) | 0.76 (0.06) | 0.75 (0.07) | 0.75 (0.07) |
| Missing | 149197 |  |  |  |  |
|  |  | N=85994 | N=31736 | N=89790 | N=17285 |
| **Heel bone mineral density** |  |  |  |  |  |
| Mean (SD) | 0.54 (0.14) | 0.52 (0.12) | 0.52 (0.12) | 0.58 (0.15) | 0.57 (0.15) |
| Missing | 180831 |  |  |  |  |
|  |  | N=38592 | N=25073 | N=42889 | N=14289 |
| **Arterial stiffness** |  |  |  |  |  |
| Mean (SD) | 9.34 (4.05) | 8.70 (4.41) | 8.71 (3.27) | 9.91 (4.27) | 9.97 (3.30) |
| Missing | 332721 |  |  |  |  |

Note: SD = standard deviation; FEV<sub>1</sub> = forced expiratory volume in one second; FVC = forced vital capacity.

### Sex differences

We found moderate to large sex differences across all physiological measures. Female participants, on average, had lower hand-grip strength, systolic and diastolic blood pressure, BMI, fat-free mass, waist-hip ratio, peak expiratory flow, FEV_1_, forced vital capacity, heel bone mineral density and arterial stiffness than males. Their pulse rate, body fat percentage, fat mass and FEV_1_/FVC ratio were higher than in males (Table 2).

**Table 2.** Sex differences in physiological measures (females compared to males)

|  |  |  |  |
| --- | --- | --- | --- |
| Variable | SMD | 95% CI |  |
| Hand-grip strength | -2.051 | -2.060 | -2.043 |
| Systolic blood pressure | -0.358 | -0.364 | -0.351 |
| Diastolic blood pressure | -0.361 | -0.368 | -0.354 |
| Pulse rate | 0.179 | 0.172 | 0.186 |
| Body mass index | -0.185 | -0.191 | -0.178 |
| Body fat percentage | 1.771 | 1.763 | 1.779 |
| Fat mass | 0.502 | 0.496 | 0.509 |
| Fat-free mass | -3.006 | -3.016 | -2.997 |
| Waist-hip ratio | -1.775 | -1.782 | -1.767 |
| Peak expiratory flow | -1.569 | -1.582 | -1.557 |
| Forced expiratory volume 1s | -1.460 | -1.473 | -1.448 |
| Forced vital capacity | -1.674 | -1.687 | -1.661 |
| FEV<sub>1</sub> / FVC | 0.165 | 0.154 | 0.176 |
| Heel bone mineral density | -0.397 | -0.405 | -0.388 |
| Arterial stiffness | -0.304 | -0.315 | -0.292 |

*Note:* SMD = standardised mean difference; CI = confidence interval; FEV<sub>1</sub> = forced expiratory volume in one second; FVC = forced vital capacity. All Bonferroni-adjusted *p*-values for Welch's *t*-test < 0.001. Negative values correspond to lower values in females.

### Case-control differences

We also found statistically significant differences between individuals with lifetime depression and healthy controls for most measures. Females with lifetime depression, on average, had lower hand-grip strength and lower blood pressure. Their pulse rate, BMI, body fat percentage, fat mass, fat-free mass, waist-hip ratio, peak expiratory flow, FEV_1_, forced vital capacity and FEV_1_/FVC ratio were higher compared to healthy controls. We did not find evidence of differences in heel bone mineral density or arterial stiffness between female cases and controls. Males with lifetime depression had lower hand-grip strength, blood pressure, peak expiratory flow, FEV_1_, forced vital capacity and heel bone mineral density, while their pulse rate, BMI, body fat percentage, fat mass, fat-free mass and waist-hip ratio were higher. We did not find evidence that the FEV_1_/FVC ratio or arterial stiffness differed between males with lifetime depression and healthy controls. The differences between individuals with lifetime depression and healthy controls were small, with the largest effect size corresponding to a standardised mean difference of 0.16 (95% CI 0.15-0.17) for fat mass in females (Table 3).

**Table 3.** Differences in physiological measures between individuals with lifetime depression and healthy controls

| Variable | Female |  |  |  |  | Male |  |  |  |  |
| --- | --- | --- | --- | --- | --- | --- | --- | --- | --- | --- |
|  | SMD | 95% CI |  | *p*<sub>Bonf.</sub> | *p*<sub>BH</sub> | SMD | 95% CI |  | *p*<sub>Bonf.</sub> | *p*<sub>BH</sub> |
| Hand-grip strength | -0.056 | -0.066 | -0.046 | <0.001 | <0.001 | -0.106 | -0.118 | -0.093 | <0.001 | <0.001 |
| Systolic blood pressure | -0.145 | -0.155 | -0.135 | <0.001 | <0.001 | -0.121 | -0.133 | -0.108 | <0.001 | <0.001 |
| Diastolic blood pressure | -0.037 | -0.047 | -0.028 | <0.001 | <0.001 | -0.041 | -0.053 | -0.028 | <0.001 | <0.001 |
| Pulse rate | 0.022 | 0.012 | 0.032 | <0.001 | <0.001 | 0.083 | 0.070 | 0.095 | <0.001 | <0.001 |
| Body mass index | 0.144 | 0.134 | 0.154 | <0.001 | <0.001 | 0.112 | 0.100 | 0.124 | <0.001 | <0.001 |
| Body fat percentage | 0.129 | 0.119 | 0.139 | <0.001 | <0.001 | 0.111 | 0.098 | 0.123 | <0.001 | <0.001 |
| Fat mass | 0.156 | 0.146 | 0.166 | <0.001 | <0.001 | 0.129 | 0.117 | 0.141 | <0.001 | <0.001 |
| Fat-free mass | 0.124 | 0.114 | 0.134 | <0.001 | <0.001 | 0.066 | 0.053 | 0.078 | <0.001 | <0.001 |
| Waist-hip ratio | 0.090 | 0.080 | 0.100 | <0.001 | <0.001 | 0.138 | 0.126 | 0.151 | <0.001 | <0.001 |
| Peak expiratory flow | 0.055 | 0.038 | 0.072 | <0.001 | <0.001 | -0.023 | -0.042 | -0.003 | 0.382 | 0.032 |
| Forced expiratory volume 1s | 0.051 | 0.034 | 0.068 | <0.001 | <0.001 | -0.024 | -0.044 | -0.005 | 0.241 | 0.022 |
| Forced vital capacity | 0.046 | 0.029 | 0.063 | <0.001 | <0.001 | -0.022 | -0.042 | -0.003 | 0.421 | 0.032 |
| FEV<sub>1</sub> / FVC | 0.018 | 0.001 | 0.035 | 0.569 | 0.044 | -0.011 | -0.031 | 0.009 | >0.999 | 0.276 |
| Heel bone mineral density | 0.013 | -0.0001 | 0.026 | 0.789 | 0.056 | -0.077 | -0.093 | -0.061 | <0.001 | <0.001 |
| Arterial stiffness | 0.004 | -0.012 | 0.020 | >0.999 | 0.571 | 0.015 | -0.004 | 0.034 | >0.999 | 0.078 |

Note: SMD = standardised mean difference; CI = confidence interval; Bonf. = Bonferroni; BH = Benjamini & Hochberg; FEV<sub>1</sub> = forced expiratory volume in one second; FVC = forced vital capacity. *P*-values for Welch's *t*-test. Negative values correspond to lower values in depression cases.

### Age-related physiological changes

Results from the unadjusted GAMs suggested that hand-grip strength, fat-free mass, lung function and heel bone mineral density declined with age, while systolic blood pressure, body fat percentage, fat mass, waist-hip ratio and arterial stiffness increased with age. Non-linear associations with age were most evident for diastolic blood pressure, pulse rate and BMI. For example, diastolic blood pressure increased until about age 55 and decreased thereafter. BMI increased with age until participants were in their mid to late 50s and plateaued thereafter, except in healthy females where BMI increased until age 70 (Figure 2-3).

**Figure 2.**
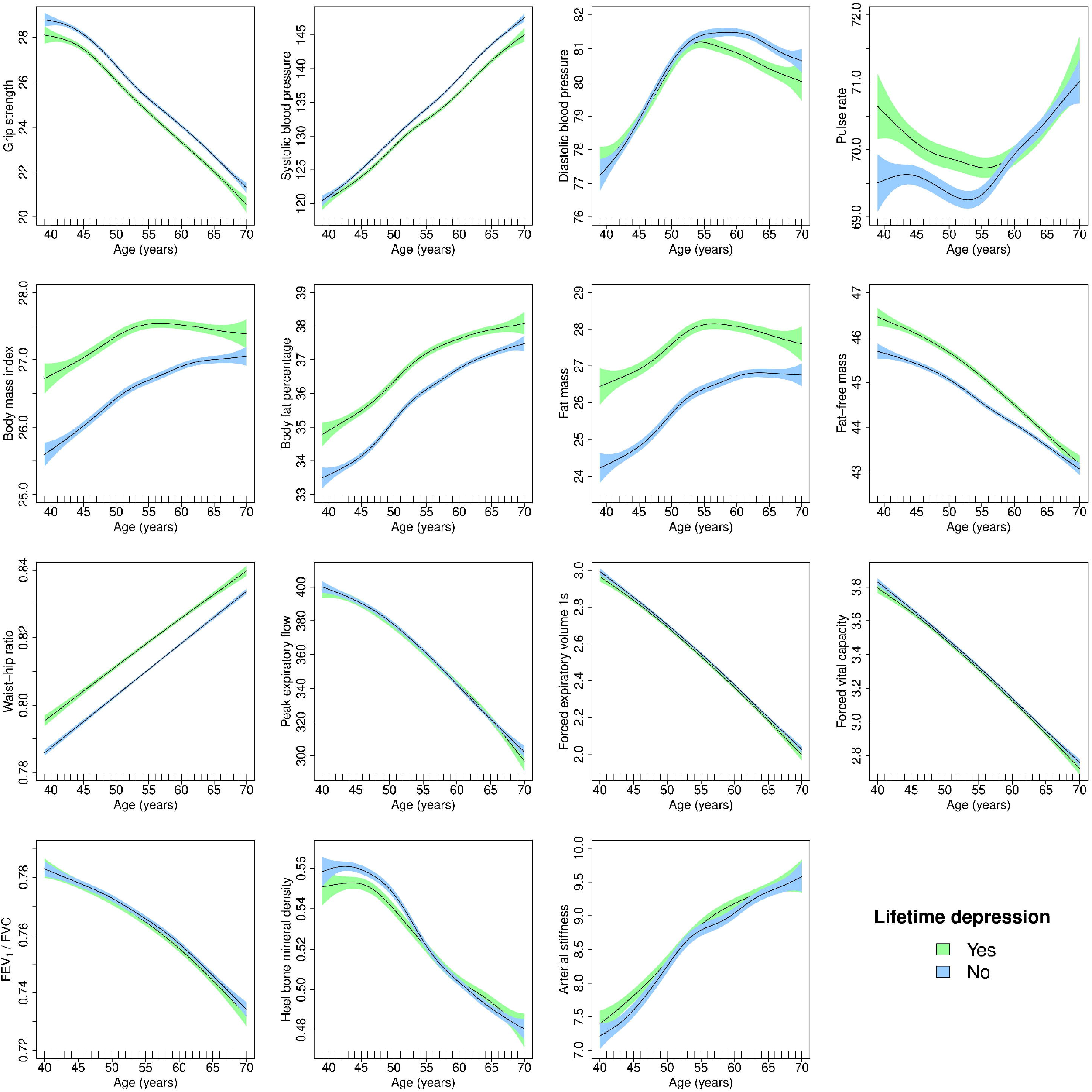
Generalised additive models of age-related changes in physiological measures in females with lifetime depression and healthy controls. The solid lines represent physiological measures against smoothing functions of age. The shaded areas correspond to approximate 95% confidence intervals (± 2 × standard error). FEV_1_ = forced expiratory volume in one second; FVC = forced vital capacity.

**Figure 3.**
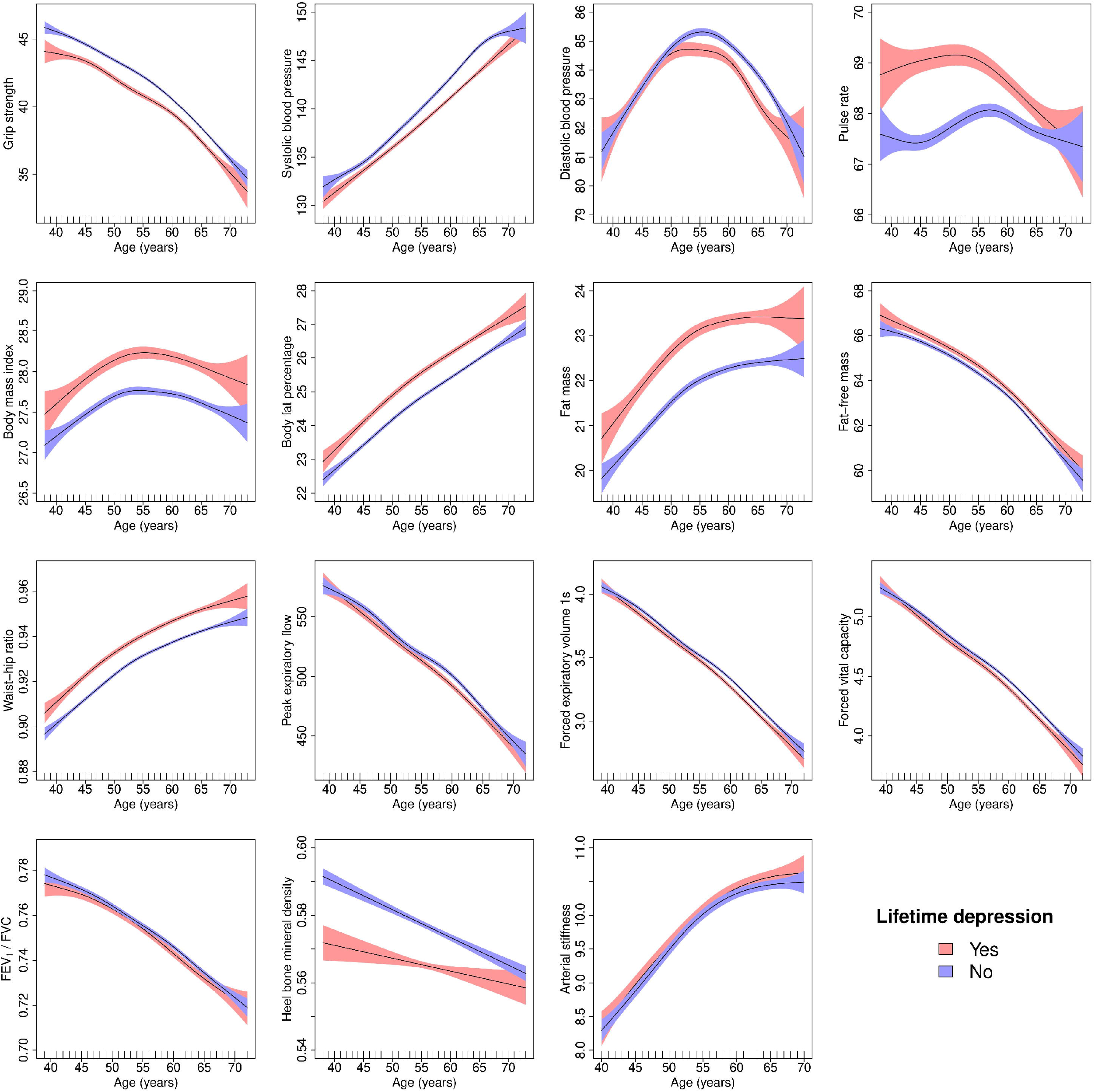
Generalised additive models of age-related changes in physiological measures in males with lifetime depression and healthy controls. The solid lines represent physiological measures against smoothing functions of age. The shaded areas correspond to approximate 95% confidence intervals (± 2 × standard error). FEV_1_ = forced expiratory volume in one second; FVC = forced vital capacity.

### Case-control differences in age-related physiological changes

Age-related changes in blood pressure, pulse rate, body composition and heel bone mineral density followed distinct trajectories in females with lifetime depression and healthy controls (Figure 4). There was also evidence of differences in age-related changes hand-in grip strength, blood pressure, pulse rate, lung function and heel bone mineral density between males with lifetime depression and healthy controls (Figure 5).

**Figure 4.**
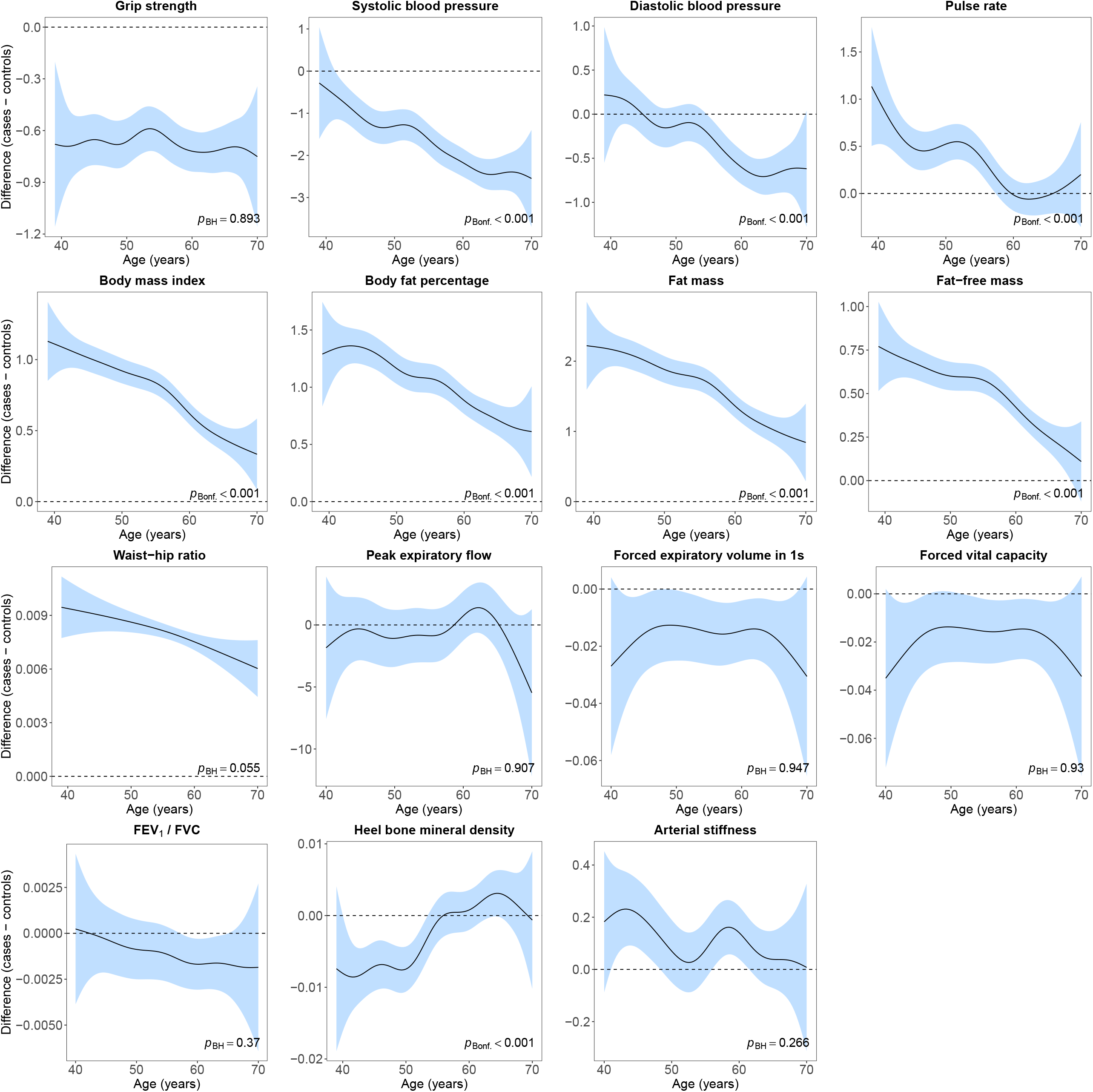
Difference smooths comparing age-related changes in physiological measures of females with lifetime depression to healthy controls. The shaded areas correspond to approximate 95% confidence intervals (± 2 × standard error). Negative values on the y-axes correspond to lower values in females with lifetime depression compared to healthy controls. The horizontal lines represent no difference between female cases and controls. FEV_1_ = forced expiratory volume in one second; FVC = forced vital capacity; Bonf. = Bonferroni; BH = Benjamini & Hochberg.

**Figure 5.**
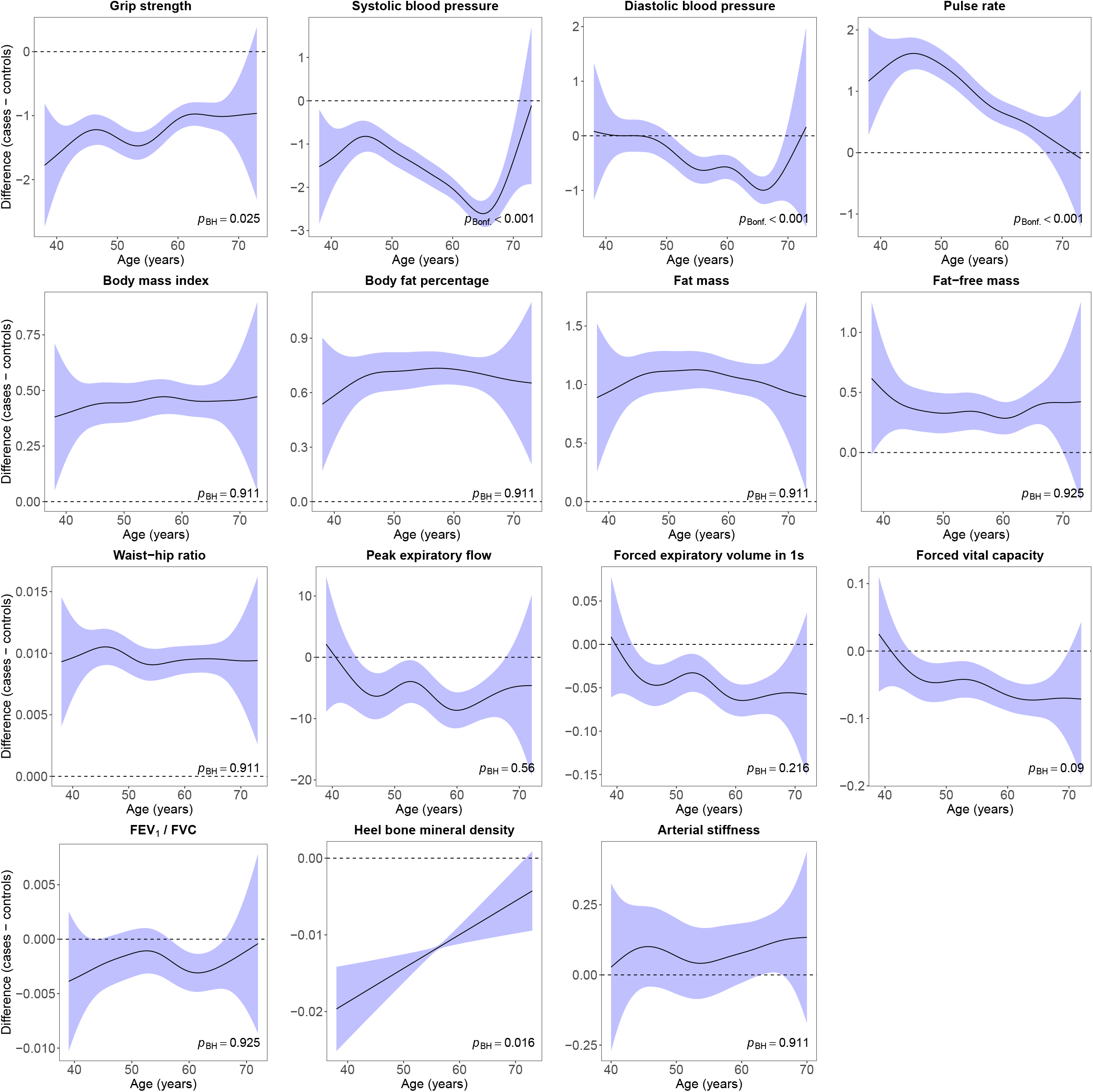
Difference smooths comparing age-related changes in physiological measures of males with lifetime depression to healthy controls. The shaded areas correspond to approximate 95% confidence intervals (± 2 × standard error). Negative values on the y-axes correspond to lower values in males with lifetime depression compared to healthy controls. The horizontal lines represent no difference between male cases and controls. FEV_1_ = forced expiratory volume in one second; FVC = forced vital capacity; Bonf. = Bonferroni; BH = Benjamini & Hochberg.

In females, we obtained similar results after adjustment for covariates, although differences between cases and controls were attenuated and there was greater overlap in the trajectories of pulse rate, lung function and heel bone mineral density. We found no evidence that age-related changes in arterial stiffness differed between female cases and controls (Supplement 7A). We also obtained similar results after adjustment for covariates in males, with some attenuation of differences between cases and controls (Supplement 7B). The formal statistical tests for differences in age-related changes in hand-grip strength and heel bone mineral density were no longer statistically significant, although age-related changes in forced vital capacity and FEV_1_ differed between male cases and controls.

### Sensitivity analyses

Case-control numbers for the sensitivity analyses are presented in Supplement 8A. We retained between 35.15% to 45.97% of cases and 100% of controls in the analyses in which cases included only those with lifetime depression according to at least two different measures. In the analyses in which we restricted our sample to participants with lifetime depression status assessed using the CIDI-SF, we retained between 24.69% to 40.67% of cases and between 22.58% to 28.03% of controls.

We found similar results when we applied more stringent criteria for lifetime depression. Average differences between female cases and controls were slightly larger for all physiological measures. In males, differences were also larger, except that there was no evidence of differences in lung function between cases and controls (Supplement 8B). Most results were also similar when we restricted our analyses to individuals with lifetime depression status assessed using the CIDI-SF. In females, differences were slightly larger, except for hand-grip strength, and there was no evidence of differences in diastolic blood pressure. In males, there was no evidence that hand-grip strength or diastolic blood pressure differed between cases and controls, and differences in lung function measures were reversed (although there was no evidence of differences in peak expiratory flow or the FEV_1_/FVC ratio). Average differences in body composition, systolic blood pressure and pulse rate were slightly larger, while differences in heel bone mineral density were slightly smaller (Supplement 8B).

Differences in age-related changes in physiological measures between female cases and controls were similar to those observed in our main analyses (Supplement 8C). There was less evidence that age-related changes in systolic blood pressure differed between males with lifetime depression according to CIDI-SF and healthy controls. There was also some evidence that age-related changes in body composition followed slightly different trajectories in males with lifetime depression according to CIDI-SF, and there was more overlap in the lung function trajectories (Supplement 8D).

## Discussion

We used data from the UK Biobank to examine differences in 15 physiological measures, between males and females, and between lifetime depression cases and healthy controls. The large sample size enabled us to highlight subtle differences between cases and controls, with heterogenous findings from previous studies that were often limited by small sample sizes. We demonstrate that case-control differences for several physiological measures vary by age, providing additional insights into the heterogeneity in ageing trajectories observed in individuals with a lifetime history of depression.

### Principal findings

We observed moderate to large sex differences across all physiological measures. We also found small but statistically significant differences between lifetime depression cases and healthy controls for most measures. All physiological measures varied with age, and we observed several differences between the trajectories of depression cases and controls. Age-related changes in body composition differed between female cases and controls, while changes in cardiovascular function differed between cases and controls of both sexes. There was some evidence of differences in the heel bone mineral density trajectories of both sexes, although in males only prior to adjustment for covariates. We found some evidence that age-related changes in lung function differed between male cases and controls after adjustment for covariates. Differences between cases and controls did not uniformly narrow or widen with age. For example, BMI in female cases was 1.1 kg/m^2^ higher at age 40 and this difference narrowed to 0.4 kg/m^2^ at age 70. In male cases, systolic blood pressure was 1 mmHg lower at age 45 and this difference widened to 2.5 mmHg at age 65.

### Findings in the context of other studies

#### Sex differences

We confirm previous research on sex differences in hand-grip strength [10, 17-19], blood pressure [20], pulse rate [21, 22], lung function [23-25], body fat percentage [26, 27], fat-free mass [26, 27] and waist-hip ratio [28]. Our finding that BMI was higher in males confirms findings from a German study [28], although studies in US and Chinese adults found no evidence of sex differences [26, 27]. The observation that fat mass was higher in females confirms findings from Sweden [29] and the US [26], although research in Chinese adults found no evidence of sex differences [27]. Males had higher heel bone mineral density, supporting previous studies of hand [30] and heel [31] bone mineral density. Our finding that arterial stiffness was higher in males supports a review which found greater arterial stiffness in males until at least age 60 [32]. However, most studies included in another systematic review found no evidence of sex differences [33], and some studies found higher arterial stiffness in females [34-36].

#### Age-related physiological changes

Our findings confirm previous research on age-related changes in hand-grip strength [10, 18, 19], blood pressure [12, 20, 37, 38], fat mass [26, 29, 39], fat-free mass [26, 39, 40], waist-hip ratio [28], lung function [23-25, 41], bone mineral density [30, 42] and arterial stiffness [21, 34]. In males, we observed a small increase in pulse rate until age 55 and a decrease thereafter, similar to data from activity trackers in >90,000 US adults [22]. However, we found a slight decrease in pulse rate in females until their mid 50s and an increase thereafter. Although we observed some increase in BMI with age, comparable to results from Swedish cohorts [29], Chinese research found no evidence of age-related changes in BMI [27]. Our finding that body fat percentage increased with age confirms previous studies [29, 40], although we found limited evidence of a plateau or decrease before age 70 observed in previous research [26].

#### Sex differences in age-related physiological changes

We confirm previous studies on sex differences in age-related changes in hand-grip strength [17, 18], blood pressure [12, 20, 37], lung function [24, 41] and bone mineral density [30, 42]. In contrast to observations that pulse rate followed parallel ageing trajectories in both sexes [22], we found changes in pulse rate in females were mirrored by inverse changes in males.

Differences in BMI narrowed with age as BMI plateaued in males at age 50 and slightly decreased thereafter, while it increased in females until at least age 65. Greater age-related increase in BMI in females has been observed previously [29]. Body fat percentage followed similar trajectories with age in both sexes. However, research in Chinese adults suggested that body fat percentage plateaued in males over 40 and continued to increase in females [27], while results from the US showed that body fat percentage decreased in females and males over 55 and 65, respectively [26]. Although Swedish data showed slightly greater age-related increase in fat mass in females [29], we found similar trajectories in both sexes. Our finding that fat mass plateaued earlier in females confirms research in US adults [26]. However, research in Chinese adults found that fat mass remained stable in males from age 40 but continued to increase in females [27]. Our study did not support the finding that fat-free mass decreased earlier in females than in males [29]. Fat-free mass decreased in both sexes, with steeper decline in males. Our finding that waist-hip ratio increased linearly with age in females confirms observations from China [43], but we did not find that waist-hip ratio plateaued in males at age 50 and decreased thereafter. Instead, we found that waist-hip ratio continued to increase until age 70. This increase attenuated at old age, in line with German research [28]. The difference between the trajectories of males and females had an inverted U-shape, with the largest difference in waist-hip ratio between age 50 to 55.

We examined absolute changes in lung function. However, a review of longitudinal studies concluded that although there was a steeper decline in absolute lung function in males, evidence of sex differences in relative decline in lung function was limited [41]. Limited previous research examined sex differences in age-related changes in the FEV_1_/FVC ratio. We found that the difference in the FEV_1_/FVC ratio narrowed with age, with some variation at the lower and upper age range.

Finally, we found only limited evidence of a steeper age-related increase in arterial stiffness in males (in our study until the age of 50) as suggested by previous studies [21, 36], or that females over 65 had higher arterial stiffness than males [32]. Sex differences in arterial stiffness narrowed between age 60 and 70.

#### Case-control differences

Our finding that hand-grip strength was lower in individuals with lifetime depression confirms previous research [44-47]. Although several studies found stronger associations between hand-grip strength and depression in females [44, 48, 49] or no sex differences [45], we observed larger differences between male cases and controls. Results from the Northern Finland 1966 birth cohort suggested that low hand-grip strength was associated with increased depressive symptoms only in males [50], while in the Irish Longitudinal Study of Ageing higher hand-grip strength was associated with lower incident depression only in females [44].

The finding that cases had lower blood pressure confirms results from the Norwegian HUNT study [51, 52]. Although several studies found lower blood pressure in depression [51-54], other research found no evidence of differences in blood pressure [55] and a meta-analysis found higher incident hypertension in depression [56]. A 2-year follow-up study suggested that low blood pressure constitutes a risk factor for depression [57]. We confirmed previous analyses showing higher pulse rate in depression [58-60].

All body composition measures were elevated in cases. A meta-analysis of observational studies found an increased risk of obesity in depression [61], and findings from the 1958 British Birth Cohort found associations between depression and both underweight and obesity [62]. Previous studies of BMI and depression yielded mixed results, including higher BMI [63], lower BMI [64, 65], U-shaped association [66] or no association [67]. A Mendelian randomisation study suggested that BMI was a causal risk factor for depression but not vice versa [68]. Mixed results have also been observed for waist-hip ratio, including lower risk of depressive symptoms in older females with high waist-hip ratio [65] and associations between depression and high waist-hip ratio [63, 69]. An analysis of UK Biobank participants found that females with probable depression had higher waist-hip ratio than males [70], although other research found higher waist-hip ratio in males with depression but not in females [71], or no evidence of an association with depressive symptoms [64]. Previous research found no evidence of associations between depression and total fat or lean mass, although central fat mass and lean mass were lower amongst older females with depression [72]. A Mendelian randomisation study suggested that fat mass, but not fat-free mass, was a causal risk factor for depression [68]. Although we found higher body fat percentage in cases, a recent analysis found associations between depression and higher body fat percentage only in females, although participants were aged 90 years [73].

In contrast to results from the Netherlands Study of Anxiety and Depression (NESDA) which suggested that peak expiratory flow was lower in females and higher in males with depression [74], we found higher peak expiratory flow in female cases. Male cases had lower peak expiratory flow, although not across all analyses. We found similar results for FEV_1_, forced vital capacity and FEV_1_/FVC ratio. Differences between male cases and controls were usually not observed in the sensitivity analyses, except that FEV_1_ and forced vital capacity were higher in the subset of male cases classified according to CIDI-SF than in controls. Research in Germany found lower FEV_1_ in depression but no evidence of differences in forced vital capacity or FEV_1_/FVC ratio [75]. Data from a community-based cohort in Korea suggested that FEV_1_, forced vital capacity and FEV_1_/FVC ratio were lower in depression, although only FEV_1_ remained lower after adjustment for covariates [76]. However, a prospective study found that lower forced vital capacity and FEV_0.75_ were associated with more depressive symptoms at follow-up in middle aged and older males [77].

We found that heel bone mineral density was lower in male cases, consistent with most previous studies [78]. There was some evidence that heel bone mineral density was higher in female cases in sensitivity analyses, contrary to most previous studies. A meta-analysis of 14 studies showed that spine and hip bone mineral density were lower in depression, especially in females [79]. One previous study suggested that heel bone mineral density was lower in female cases below the age of 40 but not in older females [80], although another study found lower heel bone mineral density also in postmenopausal females [81]. We did not find evidence that arterial stiffness differed between cases and controls. This finding contrasts with most previous studies that found higher arterial stiffness in depression [82, 83], including a recent UK Biobank study [84].

#### Case-control differences in age-related physiological changes

Hand-grip strength differences between female cases and controls were fairly stable across age, while generally narrowing with age in males. To our knowledge, the only study that examined changes in hand-grip strength in depression longitudinally found similar rates of decline in female cases and controls, in line with our findings. Contrary to our study, the authors found no evidence that hand-grip strength decreased in male cases [85].

The HUNT study showed that prospective associations between depressive symptoms and changes in blood pressure were independent of age [51]. Findings from the Baltimore Longitudinal Study of Aging found associations between greater levels of depressive symptoms and lower diastolic blood pressure until age 45, and attenuated age-related decline thereafter in females but not in males [86]. We found that case-control differences in systolic blood pressure increased with age. There was no evidence of a difference in diastolic blood pressure until participants were in their early to mid 50s, although diastolic blood pressure was lower in cases thereafter. This difference narrowed as participants approached age 70. To our knowledge, this is the first study to show that differences in pulse rate between cases and controls narrowed with age.

This is also the first study to examine age-related changes in body composition in cases compared to controls. Across these measures, we found that differences in body composition narrowed with age in females, while generally remaining constant in males. There were some inconsistencies in the sensitivity analyses with more stringent case definition. These analyses suggested an increasing difference in body fat percentage, a flat inverted U-shape for fat mass and fat-free mass, and some variation in the difference smooth for wait-hip ratio.

Results from the NESDA suggested a steeper decrease in peak expiratory flow in male cases during a 6-year follow-up, while rates of decline were similar in female cases and controls [85]. We also found some evidence of a steeper decrease in peak expiratory flow with age in male cases, although differences between female cases and controls remained stable. Rates of decline in the NESDA were similar in younger and older participants [85], concordant with our results showing an almost linear decrease in peak expiratory flow. While one previous study found a weak negative correlation between depressive symptoms and FEV_1_ only in participants older than 50 [76], our study is, to our knowledge, the first to examine case-control differences in ageing trajectories across multiple lung function measures. We found that the difference in FEV_1_ and forced vital capacity remained stable with age in females, with some variation in sensitivity analyses, while the difference widened with age in males, although not across all analyses. We did not find clear evidence of differences in the trajectories of FEV_1_/FVC ratio.

Differences in heel bone mineral density between cases and controls narrowed with age, approaching no difference in males in their late 60s and in females in their mid 50s, with some evidence of higher bone mineral density in older females. One previous study found lower heel bone mineral density in females with depression who were younger than 40 but not in older females [80]. However, lower bone mineral density has also been found in postmenopausal females with depression [87]. A meta-analysis of 23 studies found lower bone mineral density in the spine, hip and forearm in premenopausal than in postmenopausal female cases compared to controls [88]. Our finding that differences in bone mineral density narrowed with age differs from a meta-analysis of five longitudinal studies which suggested that depression was associated with increased annual loss in hip and spine bone mineral density [79]. One study showed that elderly females with higher levels of depressive symptoms had an age-adjusted mean total hip bone mineral density decrease of 0.96% per year, compared to 0.69% in females with fewer symptoms [89]. Similar findings have been observed in males [90]. However, a prospective study of premenopausal females found no evidence of decreasing bone mineral density irrespective of depressive symptoms [91]. To our knowledge, this is the first study to examine differences in age-related changes in arterial stiffness between cases and controls. While we did not find clear evidence of different trajectories with age, there was some evidence that arterial stiffness was higher in female cases younger than age 50.

Inconsistencies between studies could result from variation in sample characteristics, differences in assessment and definition of depression, or medication use. For example, previous research found that after adjustment for depression, antidepressant use was associated with lower spine bone mineral density in females but not in males [92], or that depression was associated with lower systolic blood pressure, while antidepressant use was associated with higher systolic and diastolic blood pressure [54]. Inconsistencies could also reflect differences in prevalence of specific symptoms. For example, somatic/vegetative symptoms but not mood/cognitive symptoms were previously associated with higher waist-hip ratio [69].

Differences between cases and controls could reflect disease-related processes or poor underlying health status [93] occurring more frequently in cases. They could also result from biological mechanisms affecting depression and physiology (although a recent study did not find evidence of genetic correlations between depression and body composition [94]), or from treatment-related changes, for example lower bone mineral density due to antidepressant use [92]. Behavioural factors such as unhealthy lifestyles known to affect physiology (e.g., smoking and low levels of physical activity) might also contribute. It is likely that associations between depression and physiology are bi-directional in that depression is both cause and consequence of poor physiology [87]. Sex differences might be due to the relative frequency of different symptoms experienced by males and females or differences in prevalence of comorbidities.

### Weaknesses of the study

Our research inevitably has limitations. Although we briefly remarked on this above, our analyses provide limited insights into the mechanism underpinning our results. The cross-sectional nature of our study presents additional uncertainty about whether these findings reflect changes in physiology due to ageing or cohort-specific differences in these measures and confounding factors. For example, previous research suggested that hand-grip strength is lower in younger cohorts [19], which could result in underestimating age-related decline in our study. Research on frailty indices provided evidence of stability across birth cohorts [95] as well as higher levels of frailty in more recent cohorts [96, 97]. The finding that some physiological differences narrowed with age could result from selection bias leading to older participants with depression in our study being healthier, relative to their age group, than younger participants. Selection bias resulting in healthier older adults participating in the UK Biobank at higher rates could also result in underestimating age-related changes. Finally, some concerns remain about the validity and reliability of our definition of lifetime depression [98]. To maximise the sample size available for our main analyses, we classified cases using multiple data sources. However, these sources all have specific strengths and limitations, which have been discussed in detail elsewhere [98-100]. For example, mental disorders are likely underreported in linked health records. Self-reports might contain some level of misclassification because mental disorders are heterogenous in their presentation and have fuzzy boundaries, although participants were asked to only report diagnoses made by health professionals. To address these concerns, we conducted sensitivity analyses with more stringent case definition and found comparable results. Given that mental disorders fluctuate throughout the life course, we examined lifetime depression, which includes single episode and recurrent depression. Future studies could examine differences between single episode and recurrent depression as well as changes in physiology in relation to current psychiatric status at a more fine-scale timespan longitudinally.

### Generalisability

The overall response rate of the UK Biobank was low (5.5%) and compared with non-responders, participants were older, more often female, of higher socio-economic status and reported fewer health conditions compared with data from a nationally representative survey [101]. An empirical investigation comparing the UK Biobank with data from 18 cohort studies with conventional response rates found the direction of associations to be similar, although with differences in magnitude [102]. There are additional concerns about the representativeness of individuals with mental disorders because participation in research can be influenced by selection bias due to mental health [103]. Our findings might not generalise to younger and older populations and conclusions are limited to adults between the ages of 37 to 73. Additional studies including children and adolescents as well as the elderly are needed.

### Implications

The primary aim of this study was to examine differences in age-related physiological changes between individuals with lifetime depression and healthy controls. Understanding age-related changes in physiology in individuals with depression is of public health importance, given that variation in these measures is linked to morbidity, mortality and other functional outcomes. The differences observed between cases and controls suggest that targeted screening for physiological function in middle-aged and older adults with depression is warranted to potentially mitigate excess mortality. Future studies aimed at further disentangling aetiology are needed to inform preventative and treatment strategies. Interventions such as resistance training might help improve physiological health and reduce depressive symptoms [104], although multiple factors associated with health status should be considered [105].

## Methods

### Study population

The UK Biobank is a prospective study of >500,000 UK residents aged 37–73 at baseline, recruited between 2006–2010. Details of the study rationale and design have been reported elsewhere [106]. Briefly, individuals registered with the UK National Health Service (NHS) and living within a 25-mile (∼40 km) radius of one of 22 assessment centres were invited to participate (9,238,453 postal invitations sent). Participants provided information on their sociodemographic characteristics, health behaviours and medical history. They also underwent physical examination including height, weight and blood pressure measurement, and had blood and urine samples taken. Linked hospital inpatient records are available for most participants and primary care records are currently available for approximately half of participants. A subset of 157,366 out of 339,092 invited participants (46%) completed an online follow-up mental health questionnaire (MHQ) between 2016-2017, covering 31% of all participants.

### Exposures

Age at baseline assessment was the primary explanatory variable.

Lifetime depression was assessed as part of the MHQ using a modified version of the depression module of the Composite International Diagnostic Interview Short Form (CIDI-SF) and defined according to DSM-5 criteria for major depressive disorder (Supplement 1) [107]. To achieve maximum coverage of the UK Biobank study, we also included in our definition of lifetime depression individuals who had reported during the nurse-led interview at baseline that a doctor had told them that they had depression (field 20002), participants who reported in response to a single-item question on the MHQ that a professional (doctors, nurse or person with specialist training such as psychologist or therapist) had diagnosed them with depression (field 20544), participants with a hospital inpatient record containing an ICD-10 code for depression (F32-F33; Supplement 2), participants with a primary care record containing a Read v2 or CTV3 code for depression (see [108] for diagnostic codes and data extraction procedures), and those who were classified as individuals with probable depression according to Smith et al. [109] based on additional questions that were introduced during the later stages of the baseline assessment (Supplement 3). We excluded individuals with any record of bipolar disorder or psychosis.

Healthy controls were defined as individuals who did not meet our criteria for lifetime depression and did not have a record of other mental disorders: (i) no “schizophrenia”, “mania/bipolar disorder/manic depression”, “anxiety/panic attacks”, “obsessive compulsive disorder”, “anorexia/bulimia/other eating disorder”, “post-traumatic stress disorder” reported during the nurse-led interview at baseline (field 20002); (ii) no mental disorders reported in response to the single-item question on the MHQ (field 20544); (iii) no self-reported current psychotropic medication use at baseline (Supplement 4) [100]; (iv) no linked hospital inpatient record that contained any ICD-10 Chapter V code except organic causes or substance use (F20-F99); (v) no primary care record containing a diagnostic code for mental disorders [108]; (vi) not classified as individual with probable bipolar disorder according to Smith et al. [109]; (vi) no Patient Health Questionnaire-9 (PHQ-9) sum score of ≥5, which was assessed as part of the MHQ.

### Outcomes

We examined 15 continuous physiological measures that were obtained at the baseline assessment, including maximal hand-grip strength, systolic and diastolic resting blood pressure, resting pulse rate, body mass index (BMI), waist-hip ratio, fat mass, fat-free mass, body fat percentage, peak expiratory flow, forced vital capacity (FVC), forced expiratory volume in one second (FEV_1_), FVC/FEV_1_ ratio, heel bone mineral density and arterial stiffness.

#### Physiological measures

All physiological measures were collected by certified healthcare technicians or nurses using a direct data entry system. Participants were asked to remove any outer garments, shoes, socks or tights. The assessment lasted about 10-15 minutes.

##### Hand-grip strength

Hand-grip strength in whole kilogram force units was measured using a Jamar J00105 hydraulic hand dynamometer (measurement range 0-90 kg). Participants were asked to sit upright in a chair and place their forearms on armrests. The dynamometer handle was set to the second incremental slot or, in participants with very large hands, moved to the third slot. Participants kept their elbow adjacent to their side and bent to a 90° angle with their thumb facing upward. A maximal score was obtained from each participant’s right and left hand. We used the maximal grip strength of the participant’s self-reported dominant hand. If no data on handedness were available, we used the highest value of both hands. This variable has been used previously in UK Biobank research [110], although other studies have used the highest value of left and right hand [111] or calculated the average grip strength from both hands [9]. We used absolute units because these are the simplest to use in risk assessment. A previous UK Biobank study found no evidence of differences in mortality or disease incidence prediction when grip strength was expressed in absolute terms compared to when expressed relative to anthropometric traits (height, weight, fat-free mass and BMI) [9].

##### Body composition

Weight measurements were obtained with a Tanita BC-418 MA body composition analyser, or, in individuals with pacemaker or females who reported that they were or might be pregnant, using a manual scale. Standing height was measured using a Seca 202 height measure. Body mass index (BMI) was calculated as weight divided by standing height squared (kg/m^2^). Waist and hip circumference in cm were measured using a Wessex non-stretchable sprung tape. Waist-to-hip ratio was calculated by dividing waist circumference by hip circumference. Whole body fat mass and fat-free mass in kg and body fat percentage (measurement range 1-75%) were estimated by electrical bio-impedance.

##### Pulmonary function

Volumetric measures of lung function were quantified using breath spirometry with a Vitalograph Pneumotrac 6800 following standard procedures. Participants were asked to record two to three blows, each lasting for at least six seconds, within a period of approximately six minutes. The reproducibility of the first two blows was automatically compared and, if acceptable (defined as a ≤5% difference in forced vital capacity (FVC) and forced expiratory volume in one second (FEV_1_)), a third blow was not required. FVC in litres describes the maximum amount of air that can be exhaled when blowing out as fast as possible after a deep breath. FEV_1_ in litres describes the amount of air that can be exhaled in one second when blowing out as fast as possible. We used the derived best measure for both FVC and FEV_1_ which was the maximum value from reproducible spirograms, in line with previous research [112]. Given that these measures are affected by several factors unrelated to pulmonary function (e.g., effort and body size), we also calculated the ratio of FEV_1_ to FVC. Peak expiratory flow (PEF) in litres per minute represents a person’s maximum speed of expiration. PEF is determined by physiological factors such as lung volume and elasticity or expiratory muscle strength. It is used for monitoring asthma and diagnosing chronic obstructive pulmonary disease. We calculated the average of all available readings. Spirometry was not performed in participants who confirmed or were unsure that they had any of the following contra-indications: chest infection in the last month (i.e., influenza, bronchitis, severe cold or pneumonia); history of a detached retina; heart attack or surgery to eyes, chest or abdomen in the last three months; history of a collapsed lung; pregnancy in the 1st or 3rd trimester; currently on medication for tuberculosis.

##### Heel bone mineral density

Heel bone mineral density was estimated by quantitative ultrasound assessment of the calcaneus using a Sahara Clinical Bone Sonometer. Participants were asked to sit with their back straight and had their left heel measured first. Measurement of both heels was performed only during later stages of the baseline assessment. Measures of speed of sound in metres per second and broadband ultrasound attenuation (decibels/megahertz) were combined into a quantitative ultrasound index. From this an estimate of heel bone mineral density in grams per cm^2^ was derived based on the assumption that sound waves travel differently through denser bones.

#### Cardiovascular measures

Blood pressure, pulse rate and arterial stiffness (pulse wave velocity) were measured by trained nurses during a separate stage of the baseline assessment.

##### Blood pressure

Seated systolic and diastolic resting blood pressure in millimetres of mercury (mmHg) was measured twice using an Omron 705 IT digital blood pressure monitor (measurement range 0-255 mmHg) following standard procedures. Participants were asked to loosen or remove any restrictive clothing and put their arm on a desktop. Measurements were taken from the left upper arm or, if impractical, from the right arm. If there was a problem with the measurement, it was repeated. Three different cuff sizes were available and a Seca tape was used to determine the circumference of the midpoint of the upper arm. If the largest cuff size was too small or the digital blood pressure monitor did not produce a reading, a manual sphygmomanometer was used in conjunction with a stethoscope. A second measurement was taken at least one minute after the first measure, following the same procedure. We used an average of the two readings to reduce potential measurement error.

##### Pulse rate

Resting pulse rate in beats per minute was recorded during the blood pressure measurements using the Omron 705 IT device or, exceptionally, a manual sphygmomanometer. We used an average of the two readings to reduce potential measurement error.

##### Arterial stiffness

Arterial stiffness (vascular reactivity) is an independent predictor of cardiovascular risk and mortality that can be measured rapidly, inexpensively, and without special training [113]. Resting pulse wave velocity was measured non-invasively using finger photoplethysmography with a PulseTrace PCA2 infra-red sensor. The pulse waveform was recorded over a period of 10-15 seconds, with the sensor clipped to the end of the index finger of the non-dominant hand while the participant was sitting. If the waveform did not fill at least 2/3 of the display of the device, or did not stabilize within one minute, the measurement was repeated on a larger finger or on the thumb. The shape of the waveform is directly related to the time it takes for the pulse wave to travel through the arterial tree in the lower body and to be reflected back to the finger. The time between the peaks of the waveform (the pulse wave peak-to-peak time, i.e., the difference between the peak values of direct and reflected components) was divided by the participant’s height to obtain the arterial stiffness index in metres per second. A higher score on the index represents stiffer arteries. The method has been externally validated and is highly correlated with the gold-standard carotid-femoral pulse wave velocity [114].

### Exclusion criteria

Participants for whom their genetic sex, inferred from the relative intensity of biological markers on the Y and X chromosomes, and self-reported sex did not match were excluded. Participants with missing data for any covariates or who responded “do not know” or “prefer not to answer” were excluded from all analyses.

### Covariates

Covariates included ethnicity (White, Asian, Black, Chinese, Mixed-race or other), gross annual household income (<£18,000, £18,000–30,999, £31,000–51,999, £52,000–100,000 or >£100,000), physical activity (number of days per week spent walking, engaging in moderate-intensity physical activity and engaging in vigorous-intensity physical activity for ≥10 minutes continuously), smoking status (never, former or current), alcohol intake frequency (never, special occasions only, one to three times a month, once or twice a week, three or four times a week or daily or almost daily), sleep duration (hours per day) and, for cardiovascular measures, current use of antihypertensive medications at baseline (yes/no). See our previous publication [105] for further details on these variables.

### Statistical analyses

All analyses, except the sensitivity analyses, were pre-specified prior to inspection of the data (preregistration: osf.io/pc76g) and algorithms were tested on simulated data. Statistical analyses were conducted using R (version 3.6.0).

Sample characteristics were summarised using means and standard deviations or counts and percentages. We also present the number of individuals who met our criteria for lifetime depression and healthy control. Kernel density plots of all physiological measures stratified by lifetime depression case status are presented separately for males and females. Differences between the sexes and between cases and controls were estimated using standardised mean differences (± 95% confidence intervals).

We examined the relationship between each physiological measure and age using generalised additive models (GAMs) with the ‘mgcv’ package in R [115]. Each measure was modelled against a penalised regression spline function of age with separate smooths for individuals with lifetime depression and healthy controls. GAMs are flexible modelling approaches that allow for the relationship between an outcome variable and a continuous exposure to be represented by a non-linear smooth curve while controlling for covariates. They are particularly useful if a linear model does not capture key aspects of the relationship between variables and attempt to achieve maximum goodness-of-fit while maintaining parsimony of the fitted curve to minimize overfitting. Smoothing parameters were selected using the restricted maximum likelihood method and we used the default option of ten basis functions to represent smooth terms in each model.

Two models were fit for males and females separately:

- Unadjusted model: physiological measure ∼ lifetime depression + s(age, by lifetime depression).
- Adjusted model: physiological measure ∼ lifetime depression + s(age, by lifetime depression) + covariates (ethnicity, household income, physical activity, smoking status, alcohol intake frequency and sleep duration).

where s(age, by lifetime depression) represents the smooth function for age, stratified by lifetime depression status. For cardiovascular measures, the adjusted model additionally included self-reported antihypertensive medication use.

To formally test whether the relationship between physiological measures and age differed between individuals with lifetime depression and healthy controls, we also fit, in separate analyses, models that included a reference smooth for healthy controls and a difference smooth for individuals with lifetime depression compared to healthy controls. For these analyses, lifetime depression status was coded as an ordered factor in R. If the difference smooth differs from zero, individuals with lifetime depression and healthy controls follow a different trend with age.

Adjusted *p*-values were calculated using the p.adjust() command in R to account for multiple testing across each set of analyses of the 15 physiological measures. Two methods were used: (1) Bonferroni and (2) Benjamini & Hochberg [116], all two-tailed, with *α* = .05 and false discovery rate of 5%, respectively.

To address potential concerns about the validity and reliability of our case definition, we conducted sensitivity analyses in which we restricted the samples to (i) individuals with lifetime depression according to at least two different data sources and (ii) individuals with lifetime depression status assessed using the CIDI-SF. These sensitivity analyses were not preregistered.

## Data Availability

The data used in the present study are available to all bona fide researchers for health-related research that is in the public interest, subject to an application process and approval criteria. Study materials are publicly available online at http://www.ukbiobank.ac.uk.

## Authorship contributions

CML acquired the studentship funding, interpreted the findings and critically reviewed the manuscript. JM conceived the idea of the study, acquired the data, carried out the statistical analysis, interpreted the findings, wrote the manuscript and revised the manuscript for final submission. Both authors read and approved the final manuscript. JM had full access to all data used in this study and takes responsibility for the integrity of the data and the accuracy of the data analysis.

## Acknowledgments

This research has been conducted using data from UK Biobank, a major biomedical database. This project made use of time on Rosalind HPC, funded by Guy’s & St Thomas’ Hospital NHS Trust Biomedical Research Centre (GSTT-BRC), South London & Maudsley NHS Trust Biomedical Research Centre (SLAM-BRC), and Faculty of Natural Mathematics & Science (NMS) at King’s College London.

## Ethics

Ethical approval for the UK Biobank study has been granted by the National Information Governance Board for Health and Social Care and the NHS North West Multicentre Research Ethics Committee (11/NW/0382). No project-specific ethical approval is needed. Data access permission has been granted under UK Biobank application 45514.

## Data sharing statement

The data used are available to all bona fide researchers for health-related research that is in the public interest, subject to an application process and approval criteria. Study materials are publicly available online at http://www.ukbiobank.ac.uk.

## Supplementary material

Supplementary information is available online.

