## Supplement for "Lifetime depression and age-related changes in body composition, cardiovascular function, grip strength and lung function: sex-specific analyses in the UK Biobank"

The following material accompanies the article

### Supplement 1. CIDI-SF lifetime depression criteria

**Supplement table 1.** Case definition lifetime depression

| | Definition and UK Biobank data fields |
|---|---|
| **Lifetime depression** (case):<br>At least one core symptom of major depressive disorder, most or all of the day on most or all days for a two-week period, with at least five depressive symptoms that represent a change from usual occurring over the same timescale, with some or a lot of impairment.<br><br>No record of psychosis or bipolar disorder. | (“Ever had prolonged feelings of sadness or depression” (20446) = Yes OR “Ever had prolonged loss of interest in normal activities” (20441) = Yes)<br>AND<br>“Fraction of day affected during worst episode of depression” (20436) = Most of day or All day long<br>AND<br>“Frequency of depressed days during worst episode of depression” (20439) = “Almost every day” or “Every day”<br>AND “Impact on normal roles during worst period of depression” (20440) = “Somewhat” or “A lot”<br>AND<br>Total number of symptoms endorsed (core and others) ≥5: “Ever had prolonged feelings of sadness or depression” (core) (20446), “Ever had prolonged loss of interest in normal activities” (core) (20441), “Feelings of tiredness during worst episode of depression” (20449), “Weight change during worst episode of depression” (20536), “Did your sleep change?” (20532), “Difficulty concentrating during worst depression” (20435), “Feelings of worthlessness during worst period of depression” (20450), “Thoughts of death during worst depression” (20437)<br>AND<br>**No** self-reported psychosis or mania for “Mental health problems ever diagnosed by a professional” (20544)<br>AND<br>**No** self-reported mania/bipolar disorder/manic depression or schizophrenia for “Non-cancer illness” (20002)<br>AND<br>**No** ICD-10 code for “manic episode” or “bipolar affective disorder” (F30-F31) or “schizophrenia, schizotypal and delusional disorders” (F20-F29)<br>AND<br>**No** probable bipolar disorder (20126) according to Smith et al. (2013)<br>AND<br>**No** bipolar disorder record according to the MHQ<br>AND<br>**No** primary care record of bipolar disorder or psychosis |
| *Note:* Criteria for lifetime depression adapted from Davis et al. (2020), doi: 10.1192/bjo.2019.100. CIDI-SF = Composite International Diagnostic Interview Short Form; ICD-10 = International Classification of Diseases, Tenth Revision; MHQ = mental health questionnaire. | |

#### Supplement 2. ICD-10 codes for depression

**Depressive episode**

- F32.0 Mild depressive episode
- F32.1 Moderate depressive episode
- F32.2 Severe depressive episode without psychotic symptoms
- F32.3 Severe depressive episode with psychotic symptoms
- F32.8 Other depressive episodes
- F32.9 Depressive episode, unspecified

**Recurrent depressive disorder**

- F33.0 Recurrent depressive disorder, current episode mild
- F33.1 Recurrent depressive disorder, current episode moderate
- F33.2 Recurrent depressive disorder, current episode severe without psychotic symptoms
- F33.3 Recurrent depressive disorder, current episode severe with psychotic symptoms
- F33.4 Recurrent depressive disorder, currently in remission
- F33.8 Other recurrent depressive disorders
- F33.9 Recurrent depressive disorder, unspecified

Supplement 3. Smith et al. depression criteria

**Depression** (adapted from Smith et al. (2013), doi: 10.1371/journal.pone.0075362).

**3. Single probable episode of major depression:**
4598 ever depressed/down for a whole week; plus 4609 at least two weeks duration; plus
4620 only one episode, plus 2090 ever seen a GP or 2100 a psychiatrist for nerves, anxiety, depression
OR
4631 ever anhedonic (unenthusiasm/uninterest) for a whole week; plus 5375 at least two weeks; plus
5386 only one episode; plus 2090 ever seen a GP or 2100 a psychiatrist for nerves, anxiety, depression.

**4. Probable recurrent major depression (moderate):**
4598 ever depressed/down for a whole week; plus 4609 at least two weeks duration; plus 4620 at least two episodes; plus 2090 ever seen a GP (but not a psychiatrist) for nerves, anxiety, depression
OR
4631 ever anhedonic (unenthusiasm/uninterest) for a whole week; plus 5375 at least two weeks; plus
5386 at least two episodes; plus 2090 ever seen a GP (but not a psychiatrist) for nerves, anxiety, depression.

**5. Probable recurrent major depression (severe):**
4598 ever depressed/down for a whole week; plus 4609 at least two weeks duration; plus 4620 at least two episodes; plus 2100 ever seen a psychiatrist for nerves, anxiety, depression
OR
4631 ever anhedonic (unenthusiasm/uninterest) for a whole week; plus 5375 at least two weeks; plus
5386 at least two episodes; plus 2100 ever seen a psychiatrist for nerves, anxiety, depression.

#### Supplement 4. Psychotropic medication codes

**Supplement table 2.** Psychotropic medication codes

| UK Biobank code | Drug name |
|---|---|
| 1140879616 | Amitriptyline |
| 1140921600 | Citalopram |
| 1140879540 | Fluoxetine |
| 1140867878 | Sertraline |
| 1140916282 | Venlafaxine |
| 1140909806 | Dosulepin |
| 1140867888 | Paroxetine |
| 1141152732 | Mirtazapine |
| 1141180212 | Escitalopram |
| 1140879634 | Trazodone |
| 1140867876 | Prozac |
| 1140882236 | Seroxat |
| 1141190158 | Cipralex |
| 1141200564 | Duloxetine |
| 1140867726 | Lofepramine |
| 1140879620 | Clomipramine |
| 1140867818 | Nortriptyline |
| 1140879630 | Imipramine |
| 1140879628 | Dothiepin |
| 1141151946 | Cipramil |
| 1140867948 | Amitriptyline |
| 1140867624 | Prothiaden |
| 1140867756 | Trimipramine |
| 1140867884 | Lustral |
| 1141151978 | Reboxetine |
| 1141152736 | Zispin |
| 1141201834 | Cymbalta |
| 1140867690 | Anafranil |
| 1140867640 | Doxepin |
| 1140867920 | Moclobemide |
| 1140867850 | Phenelzine |
| 1140879544 | Fluvoxamine |
| 1141200570 | Yentreve |
| 1140867934 | Triptafen |
| 1140867758 | Surmontil |
| 1140867914 | Tranylcypromine |
| 1140867820 | Allegron |
| 1141151982 | Edronax |
| 1140882244 | Molipaxin |
| 1140879556 | Mianserin |
| 1140867852 | Nardil |
| 1140867860 | Faverin |
| 1140917460 | Nefazodone |
| 1140867938 | Amitriptyline+Chlordiazepoxide |
| 1140867856 | Isocarboxazid |
| 1140867922 | Manerix |
| 1140910820 | Maoi |
| 1140882312 | Sinequan |
| 1140867944 | Tranylcypromine+Trifluoperazine |
| 1140867784 | Ludiomil |
| 1140867812 | Norval |
| 1140867668 | Tryptizol |
| 1140867940* | Fluphenazine hydrochloride+Nortriptyline 1.5mg/30mg tablet |
| 1140867942* | Fluphenazine hcl+Nortriptyline 500micrograms/10mg tablet |
| 1140928916 | Olanzapine |
| 1141152848 | Quetiapine |
| 1140867444 | Risperidone |
| 1140879658 | Chlorpromazine |
| 1140868120 | Trifluoperazine |
| 1141153490 | Amisulpride |
| 1140867304 | Sulpiride |
| 1141152860 | Seroquel |
| 1140867168 | Haloperidol |
| 1141195974 | Aripiprazole |
| 1140867244 | Stelazine |
| 1140867152 | Depixol |
| 1140909800 | Flupentixol |
| 1140867420 | Clozapine |
| 1140879746 | Promazine |
| 1141177762 | Risperdal |
| 1140867456 | Modecate |
| 1140867952 | Fluanxol |
| 1140867150 | Flupenthixol |
| 1141167976 | Zyprexa |

| | |
|---|---|
| 1140882100 | Zuclopenthixol |
| 1140867342 | Clopixol |
| 1140863416 | Largactil |
| 1141202024 | Abilify |
| 1140882098 | Fluphenazine |
| 1140867184 | Haldol |
| 1140867092 | Serenace |
| 1140882320 | Clozaril |
| 1140910358 | Chlorpromazine |
| 1140867208 | Perphenazine |
| 1140909802 | Levomepromazine |
| 1140867134 | Pericyazine |
| 1140867306 | Dolmatil |
| 1140867210 | Fentazin |
| 1140867398 | Fluphenazine |
| 1140867078 | Benperidol |
| 1140867218 | Pimozide |
| 1141201792 | Zaponex |
| 1141200458 | Denzapine |
| 1140867136 | Neulactil |
| 1140879750 | Thioridazine |
| 1140867180 | Dozic |
| 1140867546 | Fluspirilene |
| 1140928260 | Panadeine |
| 1140927956 | Sertindole |
| 1140867490 | Lithium product |
| 1140867494 | Camcolit 250 tablet |
| 1140867498 | Liskonum 450mg m/r tablet |
| 1140867500 | Phasal 300mg m/r tablet |
| 1140867504 | Priadel 200mg m/r tablet |
| 1140867518 | Litarex 564mg m/r tablet |
| 1140867520 | Li-liquid 5.4mmol/5ml oral solution |

*Note:* Adapted from Davis et al. (2019), doi: 10.1002/mpr.1796. *medication code not included in Davis et al. (2019).

#### Supplement 5. Density plots physiological measures

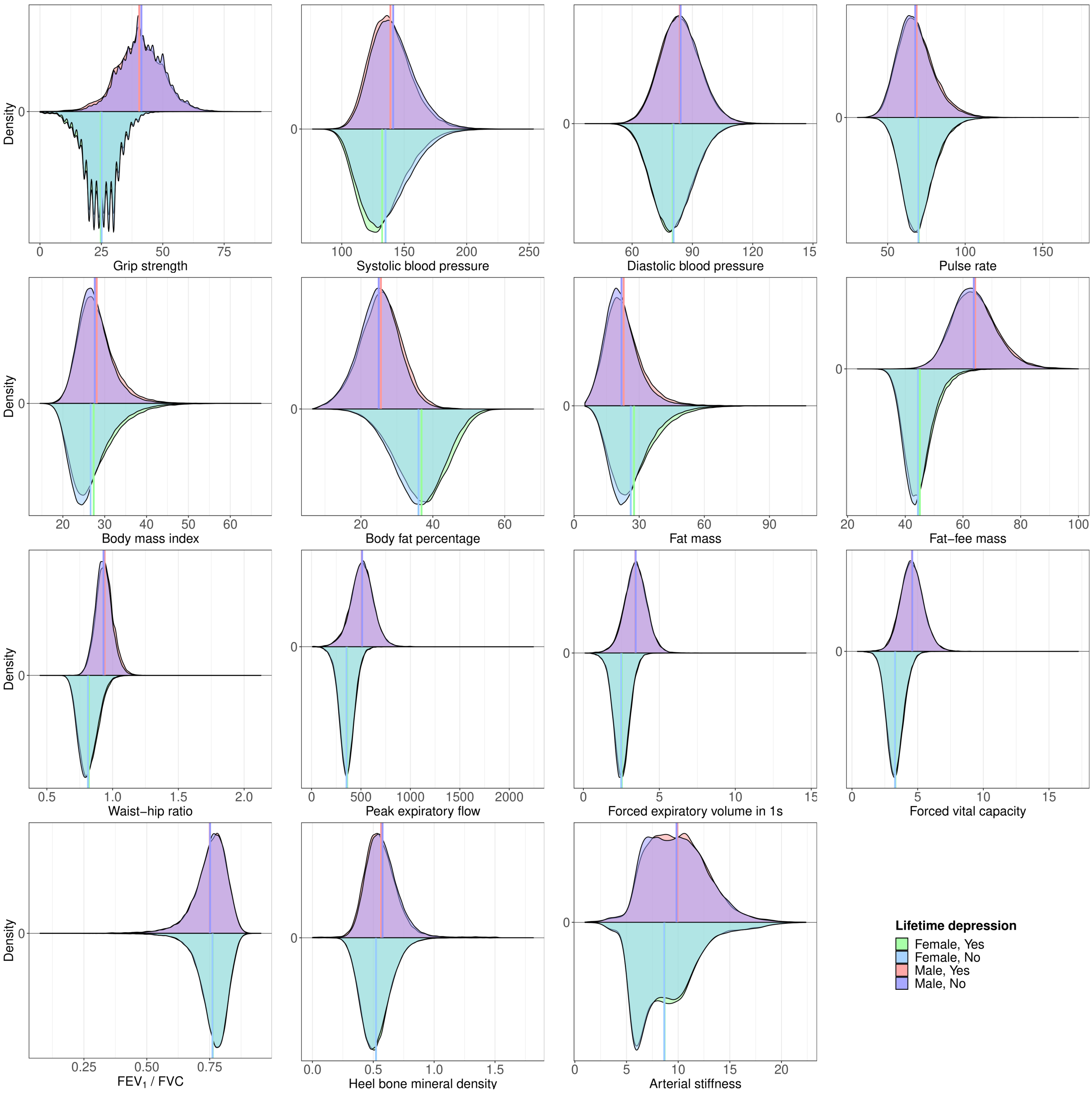

**Supplement figure 1.** Physiological measures of males and females with lifetime depression and healthy controls. Arterial stiffness truncated at 99.9th %ile. $FEV_1$ = forced expiratory volume in one second; FVC = forced vital capacity.

### Supplement 6. Sample characteristics

**Supplement table 3.** Sample characteristics

| | | Female | | Male | |
|---|---|---|---|---|---|
| | Overall (N=502521) | Healthy control (N=123842) | Depression (N=56276) | Healthy control (N=131212) | Depression (N=31063) |
| **Age** | | | | | |
| Mean (SD) | 56.53 (8.10) | 55.90 (8.03) | 54.74 (7.78) | 56.37 (8.26) | 55.71 (7.96) |
| **Ethnicity** | | | | | |
| White | 472711 (94.1%) | 117587 (94.9%) | 54386 (96.6%) | 125053 (95.3%) | 30089 (96.9%) |
| Mixed-race | 2958 (0.6%) | 805 (0.7%) | 434 (0.8%) | 588 (0.4%) | 145 (0.5%) |
| Asian | 8061 (1.6%) | 2096 (1.7%) | 556 (1.0%) | 1779 (1.4%) | 195 (0.6%) |
| Black | 9882 (2.0%) | 1809 (1.5%) | 459 (0.8%) | 2518 (1.9%) | 393 (1.3%) |
| Chinese | 1574 (0.3%) | 527 (0.4%) | 91 (0.2%) | 357 (0.3%) | 37 (0.1%) |
| Other | 4558 (0.9%) | 1018 (0.8%) | 350 (0.6%) | 917 (0.7%) | 204 (0.7%) |
| Prefer not to answer | 1662 (0.3%) | | | | |
| Do not know | 217 (<0.1%) | | | | |
| Missing | 898 (0.2%) | | | | |
| **Household income**[1] | | | | | |
| Very low | 97205 (19.3%) | 26243 (21.2%) | 13986 (24.9%) | 21903 (16.7%) | 7181 (23.1%) |
| Low | 108177 (21.5%) | 32464 (26.2%) | 14751 (26.2%) | 31452 (24.0%) | 7472 (24.1%) |
| Middle | 110774 (22.0%) | 32735 (26.4%) | 14575 (25.9%) | 36631 (27.9%) | 8233 (26.5%) |
| High | 86269 (17.2%) | 25352 (20.5%) | 10605 (18.8%) | 31999 (24.4%) | 6599 (21.2%) |
| Very high | 22930 (4.6%) | 7048 (5.7%) | 2359 (4.2%) | 9227 (7.0%) | 1578 (5.1%) |
| Prefer not to answer | 49848 (9.9%) | | | | |
| Do not know | 21305 (4.2%) | | | | |
| Missing | 6013 (1.2%) | | | | |
| **Walking**[2] | | | | | |
| Mean (SD) | 5.39 (1.93) | 5.48 (1.84) | 5.35 (1.95) | 5.31 (1.99) | 5.23 (2.06) |
| Prefer not to answer | 979 (0.2%) | | | | |
| Unable to walk | 1929 (0.4%) | | | | |
| Do not know | 6687 (1.3%) | | | | |
| Missing | 874 (0.2%) | | | | |
| **Moderate activity**[2] | | | | | |
| Mean (SD) | 3.63 (2.33) | 3.62 (2.31) | 3.51 (2.34) | 3.59 (2.31) | 3.46 (2.35) |
| Prefer not to answer | 2273 (0.5%) | | | | |
| Do not know | 24120 (4.8%) | | | | |
| Missing | 878 (0.2%) | | | | |
| **Vigorous activity**[2] | | | | | |
| Mean (SD) | 1.84 (1.96) | 1.73 (1.85) | 1.61 (1.82) | 2.10 (2.02) | 1.95 (2.02) |
| Prefer not to answer | 4116 (0.8%) | | | | |
| Do not know | 22582 (4.5%) | | | | |
| Missing | 878 (0.2%) | | | | |
| **Smoking status** | | | | | |
| Never | 273528 (54.4%) | 77069 (62.2%) | 30307 (53.9%) | 67927 (51.8%) | 13959 (44.9%) |
| Former | 173064 (34.4%) | 37682 (30.4%) | 19633 (34.9%) | 49044 (37.4%) | 12473 (40.2%) |
| Current | 52979 (10.5%) | 9091 (7.3%) | 6336 (11.3%) | 14241 (10.9%) | 4631 (14.9%) |
| Prefer not to answer | 2059 (0.4%) | | | | |
| Missing | 891 (0.2%) | | | | |
| **Alcohol intake frequency** | | | | | |
| Never | 40645 (8.1%) | 9217 (7.4%) | 4658 (8.3%) | 6290 (4.8%) | 2323 (7.5%) |
| Special occasions | 58011 (11.5%) | 16310 (13.2%) | 8361 (14.9%) | 8259 (6.3%) | 2436 (7.8%) |
| 1-3/month | 55856 (11.1%) | 15862 (12.8%) | 7869 (14.0%) | 11184 (8.5%) | 3054 (9.8%) |
| 1-2/week | 129294 (25.7%) | 33072 (26.7%) | 14017 (24.9%) | 34139 (26.0%) | 7516 (24.2%) |
| 3-4/week | 115443 (23.0%) | 28093 (22.7%) | 11628 (20.7%) | 36606 (27.9%) | 7587 (24.4%) |
| Daily/almost daily | 101770 (20.3%) | 21288 (17.2%) | 9743 (17.3%) | 34734 (26.5%) | 8147 (26.2%) |
| Prefer not to answer | 605 (0.1%) | | | | |
| Missing | 897 (0.2%) | | | | |
| **Sleep duration** | | | | | |
| Mean (SD) | 7.15 (1.11) | 7.18 (1.00) | 7.17 (1.17) | 7.13 (0.99) | 7.11 (1.21) |
| Prefer not to answer | 386 (0.1%) | | | | |
| Do not know | 2943 (0.6%) | | | | |
| Missing | 887 (0.2%) | | | | |

*Note:* Descriptive statistics for covariates based on main dataset *N*=342,393. [1]Annual household income groups: very low (<£18,000), low (£18,000–30,999), middle (£31,000–51,999), high (£52,000–100,000) and very high (>£100,000). [2]number of days per week engaging in these activities for 10+ minutes continuously.

### Supplement 7. Adjusted GAMs

#### 7A. Age-related changes in females

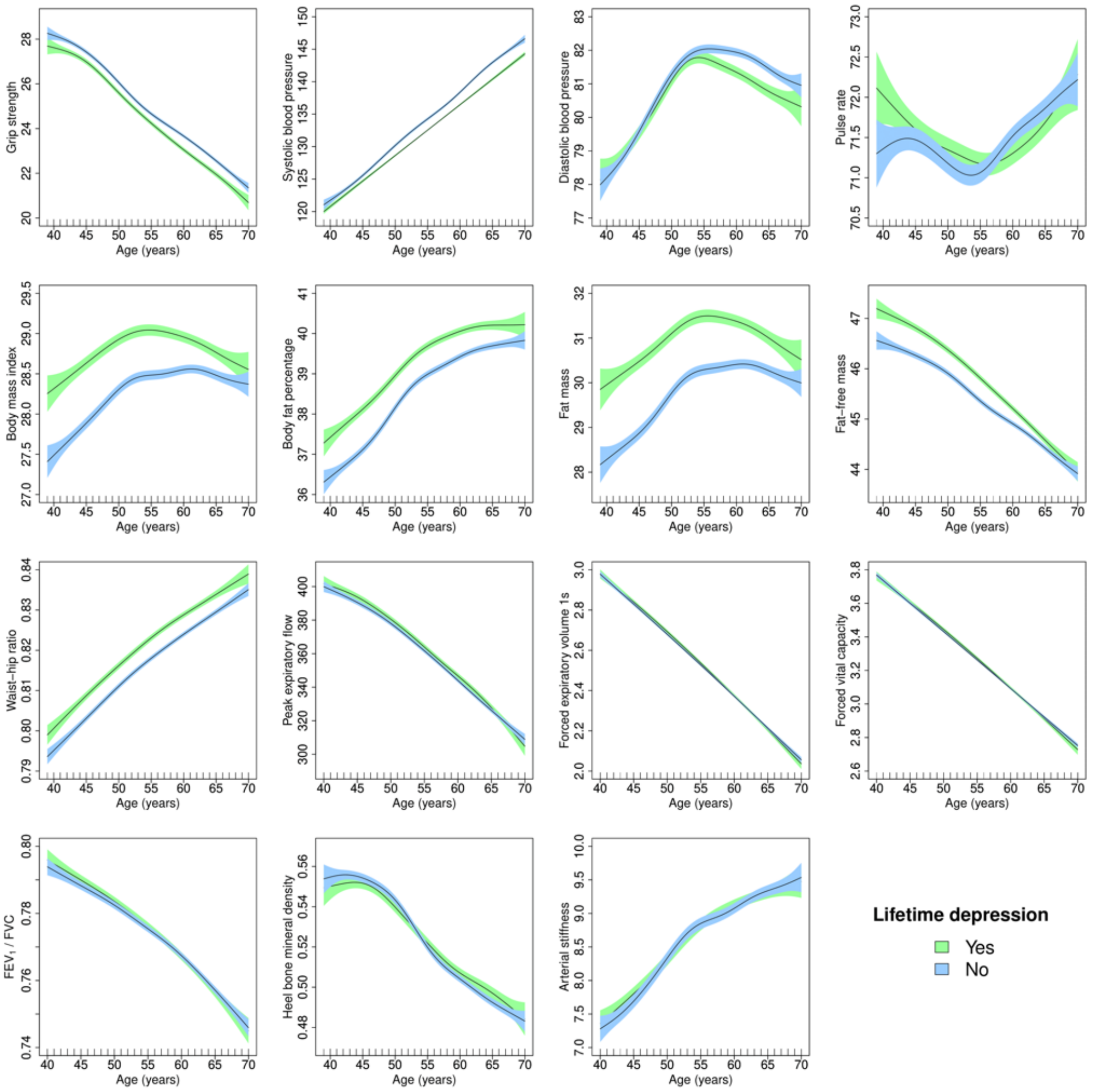

**Supplement figure 2.** Adjusted generalised additive models of age-related changes in physiological measures in females with lifetime depression and healthy controls. Models were adjusted for ethnicity (except lung function), gross annual household income, physical activity, smoking status, alcohol intake frequency, sleep duration and, for cardiovascular measures, current use of antihypertensive medications. The solid lines represent physiological measures against smoothing functions of age. The shaded areas correspond to approximate 95% confidence intervals (± 2 × standard error). $FEV_1$ = forced expiratory volume in one second; FVC = forced vital capacity.

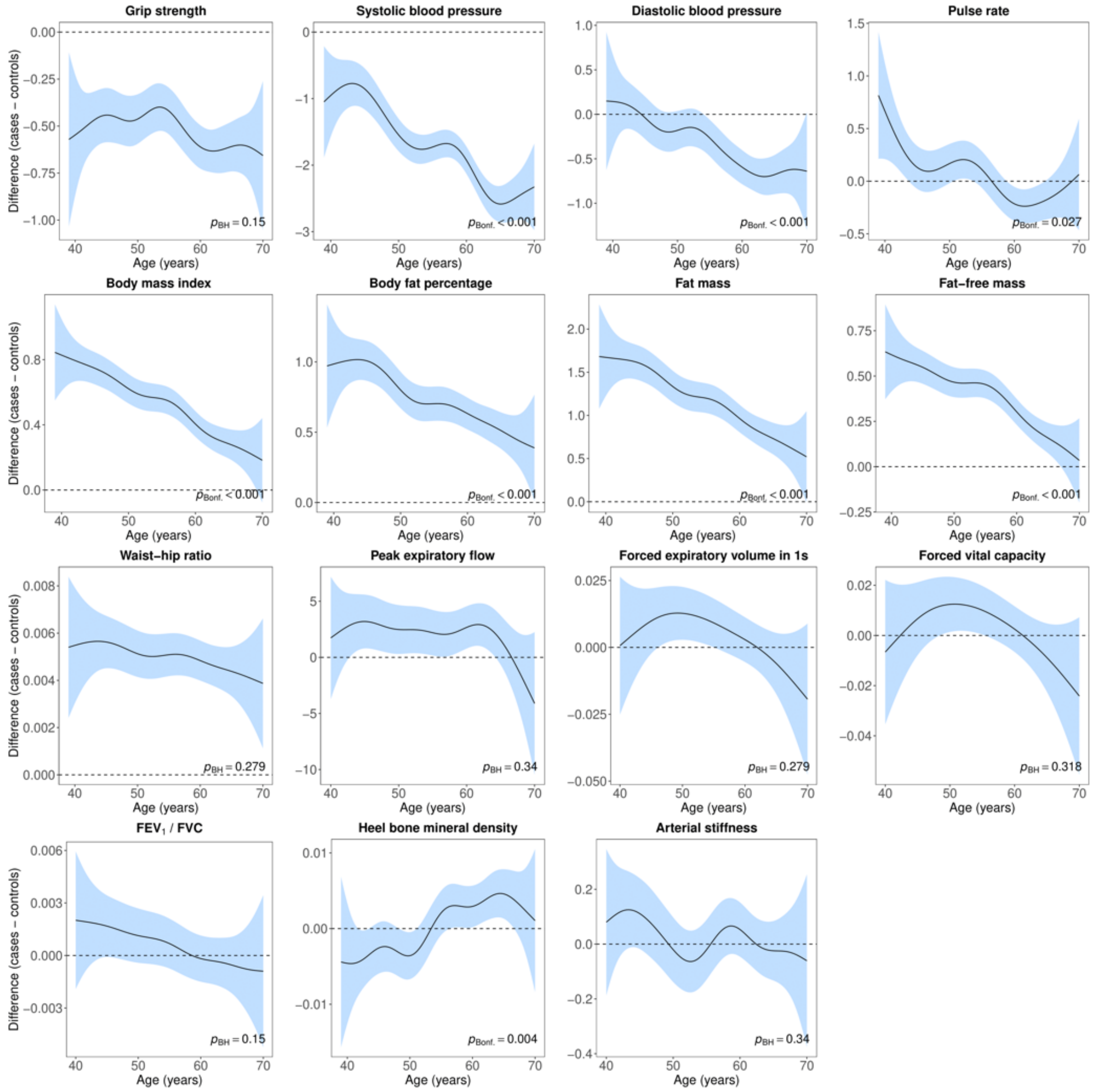

**Supplement figure 3.** Difference smooths comparing age-related changes in physiological measures of females with lifetime depression to healthy controls. Models were adjusted for ethnicity (except lung function), gross annual household income, physical activity, smoking status, alcohol intake frequency, sleep duration and, for cardiovascular measures, current use of antihypertensive medications. The shaded areas correspond to approximate 95% confidence intervals (± 2 × standard error). Negative values on the y-axes correspond to lower values in females with lifetime depression compared to healthy controls. The horizontal lines represent no difference between female cases and controls. $FEV_1$ = forced expiratory volume in one second; FVC = forced vital capacity; Bonf. = Bonferroni; BH = Benjamini & Hochberg.

7B. Age-related changes in males

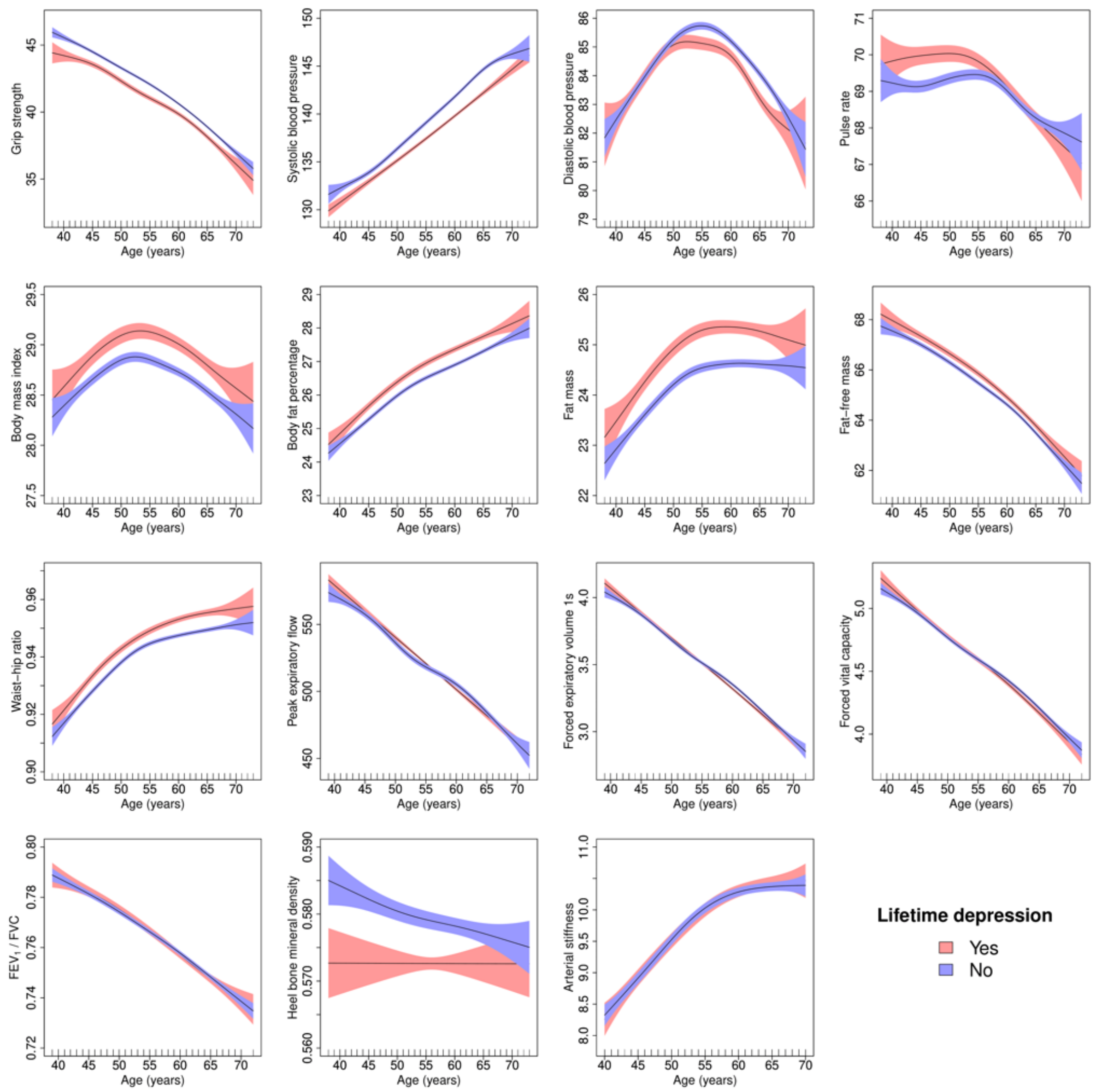

**Supplement figure 4.** Adjusted generalised additive models of age-related changes in physiological measures in males with lifetime depression and healthy controls. Models were adjusted for ethnicity (except lung function), gross annual household income, physical activity, smoking status, alcohol intake frequency, sleep duration and, for cardiovascular measures, current use of antihypertensive medications. The solid lines represent physiological measures against smoothing functions of age. The shaded areas correspond to approximate 95% confidence intervals (± 2 × standard error). $FEV_1$ = forced expiratory volume in one second; FVC = forced vital capacity.

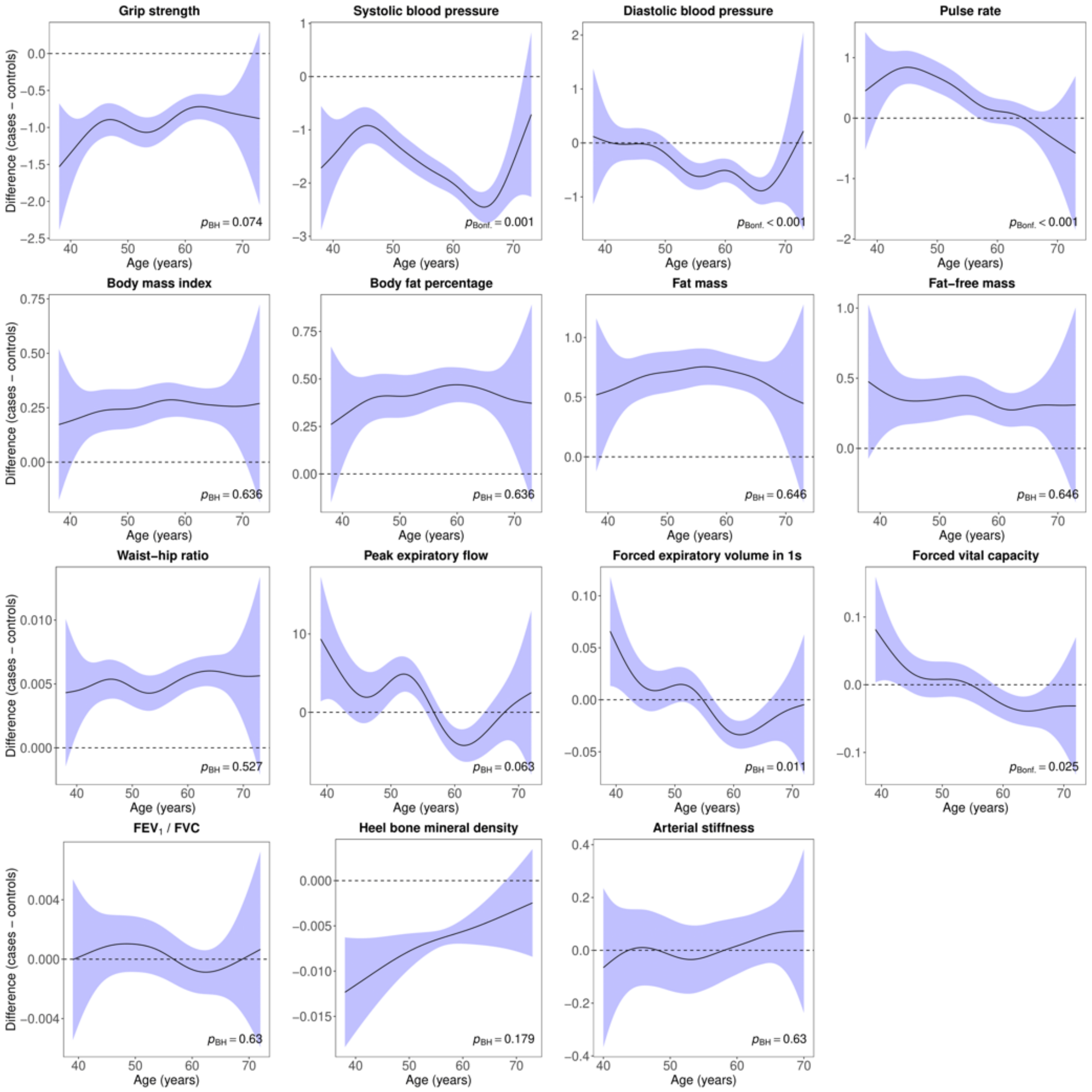

**Supplement figure 5.** Difference smooths comparing age-related changes in physiological measures of males with lifetime depression to healthy controls. Models were adjusted for ethnicity (except lung function), gross annual household income, physical activity, smoking status, alcohol intake frequency, sleep duration and, for cardiovascular measures, current use of antihypertensive medications. The shaded areas correspond to approximate 95% confidence intervals (± 2 × standard error). Negative values on the y-axes correspond to lower values in males with lifetime depression compared to healthy controls. The horizontal lines represent no difference between male cases and controls. $FEV_1$ = forced expiratory volume in one second; FVC = forced vital capacity; Bonf. = Bonferroni; BH = Benjamini & Hochberg.

### Supplement 8. Sensitivity analyses

#### 8A. Case-control numbers

**Supplement table 4.** Case-control numbers sensitivity analyses

| | | ≥ 2 depression measures | | CIDI-SF lifetime depression (MHQ) | |
|---|---|---|---|---|---|
| Dataset | Sex | Healthy control | Depression | Healthy control | Depression |
| Main dataset | Female | 123842 | 23517 | 30113 | 20482 |
| | Male | 131212 | 11785 | 32776 | 9432 |
| Lung function | Female | 43133 | 8010 | 10750 | 7162 |
| | Male | 52359 | 4617 | 13586 | 3826 |
| Bone mineral density | Female | 85994 | 12219 | 19420 | 12907 |
| | Male | 89790 | 6075 | 21289 | 6005 |
| Arterial stiffness | Female | 38592 | 11525 | 10818 | 7688 |
| | Male | 42889 | 5899 | 11805 | 3528 |

*Note:* CIDI-SF = Composite International Diagnostic Interview Short Form; MHQ = mental health questionnaire.

#### 8B. Case control differences

**Supplement table 5.** Differences in physiological measures between individuals with lifetime depression and healthy controls

| | ≥ 2 depression measures | | | | | CIDI-SF lifetime depression (MHQ) | | | | |
|---|---|---|---|---|---|---|---|---|---|---|
| Variable | SMD | 95% CI | | $p_{Bonf.}$ | $p_{BH}$ | SMD | 95% CI | | $p_{Bonf.}$ | $p_{BH}$ |
| **Female** | | | | | | | | | | |
| Hand-grip strength | -0.067 | -0.081 | -0.053 | <0.001 | <0.001 | -0.042 | -0.060 | -0.025 | <0.001 | <0.001 |
| Systolic blood pressure | -0.192 | -0.206 | -0.178 | <0.001 | <0.001 | -0.162 | -0.180 | -0.145 | <0.001 | <0.001 |
| Diastolic blood pressure | -0.047 | -0.061 | -0.033 | <0.001 | <0.001 | -0.005 | -0.023 | 0.013 | >0.999 | 0.600 |
| Pulse rate | 0.028 | 0.014 | 0.042 | 0.002 | <0.001 | 0.052 | 0.034 | 0.070 | <0.001 | <0.001 |
| Body mass index | 0.190 | 0.176 | 0.204 | <0.001 | <0.001 | 0.230 | 0.212 | 0.248 | <0.001 | <0.001 |
| Body fat percentage | 0.165 | 0.151 | 0.179 | <0.001 | <0.001 | 0.193 | 0.175 | 0.210 | <0.001 | <0.001 |
| Fat mass | 0.211 | 0.197 | 0.225 | <0.001 | <0.001 | 0.227 | 0.209 | 0.245 | <0.001 | <0.001 |
| Fat-free mass | 0.181 | 0.167 | 0.195 | <0.001 | <0.001 | 0.166 | 0.148 | 0.184 | <0.001 | <0.001 |
| Waist-hip ratio | 0.115 | 0.101 | 0.129 | <0.001 | <0.001 | 0.122 | 0.104 | 0.139 | <0.001 | <0.001 |
| Peak expiratory flow | 0.120 | 0.096 | 0.144 | <0.001 | <0.001 | 0.073 | 0.043 | 0.103 | <0.001 | <0.001 |
| Forced expiratory volume 1s | 0.110 | 0.086 | 0.134 | <0.001 | <0.001 | 0.067 | 0.037 | 0.097 | <0.001 | <0.001 |
| Forced vital capacity | 0.102 | 0.078 | 0.126 | <0.001 | <0.001 | 0.049 | 0.019 | 0.079 | 0.021 | 0.002 |
| $FEV_1$ / FVC | 0.043 | 0.019 | 0.067 | 0.006 | <0.001 | 0.051 | 0.021 | 0.081 | 0.011 | 0.001 |
| Heel bone mineral density | 0.024 | 0.005 | 0.043 | 0.177 | 0.013 | 0.028 | 0.005 | 0.050 | 0.227 | 0.017 |
| Arterial stiffness | 0.007 | -0.014 | 0.028 | >0.999 | 0.448 | 0.016 | -0.013 | 0.045 | >0.999 | 0.301 |
| **Male** | | | | | | | | | | |
| Hand-grip strength | -0.116 | -0.135 | -0.097 | <0.001 | <0.001 | -0.002 | -0.025 | 0.020 | >0.999 | 0.900 |
| Systolic blood pressure | -0.159 | -0.178 | -0.140 | <0.001 | <0.001 | -0.143 | -0.166 | -0.120 | <0.001 | <0.001 |
| Diastolic blood pressure | -0.044 | -0.063 | -0.025 | <0.001 | <0.001 | -0.002 | -0.025 | 0.021 | >0.999 | 0.900 |
| Pulse rate | 0.109 | 0.090 | 0.128 | <0.001 | <0.001 | 0.088 | 0.066 | 0.111 | <0.001 | <0.001 |
| Body mass index | 0.140 | 0.121 | 0.158 | <0.001 | <0.001 | 0.192 | 0.169 | 0.215 | <0.001 | <0.001 |
| Body fat percentage | 0.134 | 0.115 | 0.153 | <0.001 | <0.001 | 0.142 | 0.119 | 0.165 | <0.001 | <0.001 |
| Fat mass | 0.165 | 0.146 | 0.184 | <0.001 | <0.001 | 0.183 | 0.160 | 0.206 | <0.001 | <0.001 |
| Fat-free mass | 0.097 | 0.078 | 0.116 | <0.001 | <0.001 | 0.137 | 0.114 | 0.160 | <0.001 | <0.001 |
| Waist-hip ratio | 0.167 | 0.148 | 0.185 | <0.001 | <0.001 | 0.167 | 0.144 | 0.190 | <0.001 | <0.001 |
| Peak expiratory flow | 0.005 | -0.025 | 0.035 | >0.999 | 0.793 | 0.019 | -0.017 | 0.055 | >0.999 | 0.413 |
| Forced expiratory volume 1s | 0.003 | -0.027 | 0.033 | >0.999 | 0.837 | 0.045 | 0.009 | 0.081 | 0.224 | 0.022 |
| Forced vital capacity | 0.011 | -0.019 | 0.041 | >0.999 | 0.564 | 0.046 | 0.011 | 0.082 | 0.190 | 0.021 |
| $FEV_1$ / FVC | -0.013 | -0.043 | 0.017 | >0.999 | 0.517 | 0.006 | -0.030 | 0.042 | >0.999 | 0.900 |
| Heel bone mineral density | -0.101 | -0.127 | -0.075 | <0.001 | <0.001 | -0.074 | -0.102 | -0.045 | <0.001 | <0.001 |
| Arterial stiffness | 0.020 | -0.007 | 0.047 | >0.999 | 0.092 | 0.001 | -0.037 | 0.038 | >0.999 | 0.974 |

*Note:* SMD = standardised mean difference; CI = confidence interval; Bonf. = Bonferroni; BH = Benjamini & Hochberg; $FEV_1$ = forced expiratory volume in one second; FVC = forced vital capacity; CIDI-SF = Composite International Diagnostic Interview Short Form; MHQ = mental health questionnaire. *P*-values for Welch's t-test.

8C. Age-related changes in females

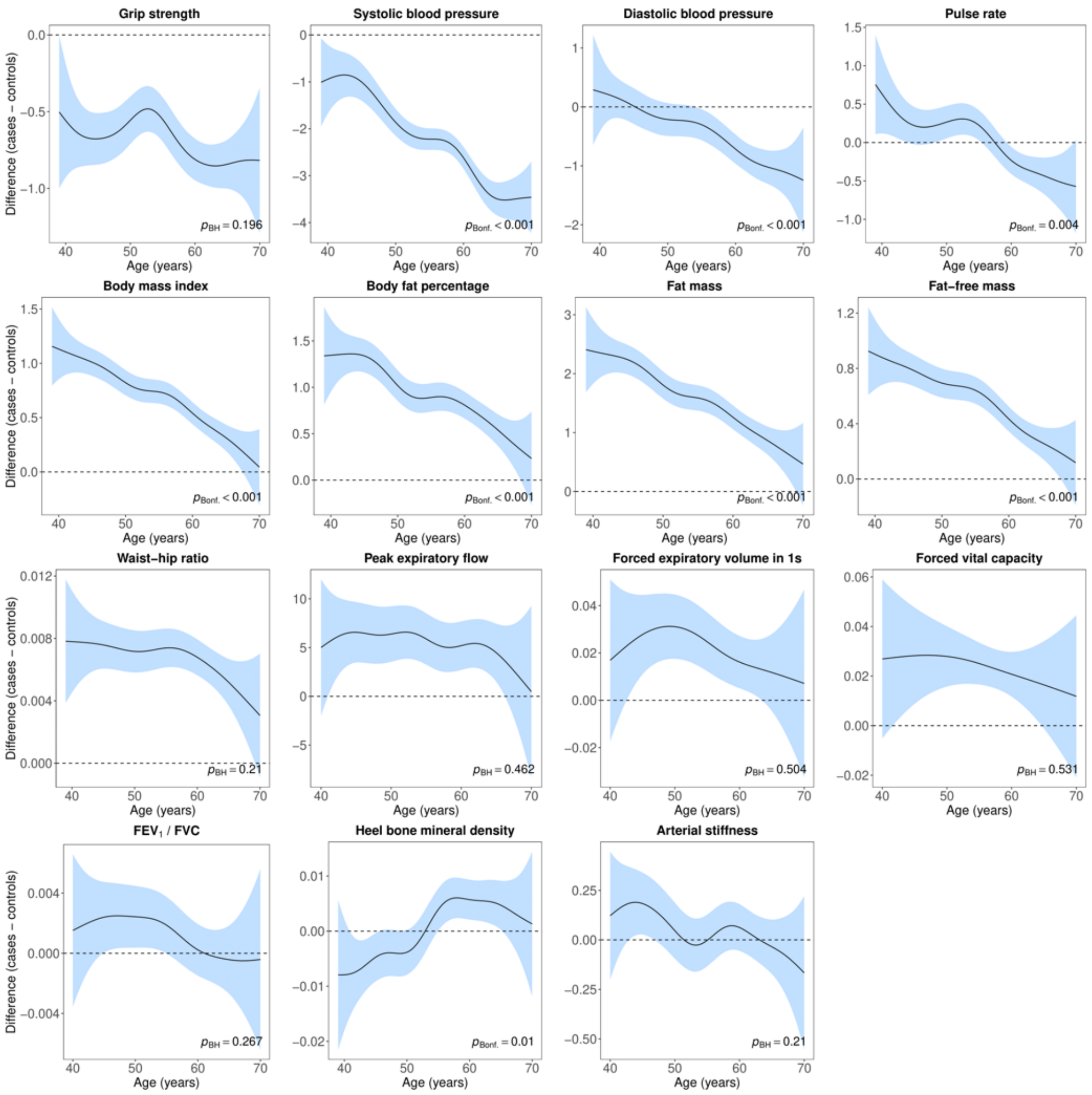

**Supplement figure 6.** Difference smooths comparing age-related changes in physiological measures of females with lifetime depression (**≥ 2 depression measures**) to healthy controls. Models were adjusted for ethnicity (except lung function), gross annual household income, physical activity, smoking status, alcohol intake frequency, sleep duration and, for cardiovascular measures, current use of antihypertensive medications. The shaded areas correspond to approximate 95% confidence intervals (± 2 × standard error). Negative values on the y-axes correspond to lower values in females with lifetime depression compared to healthy controls. The horizontal lines represent no difference between female cases and controls. $FEV_1$ = forced expiratory volume in one second; FVC = forced vital capacity; Bonf. = Bonferroni; BH = Benjamini & Hochberg.

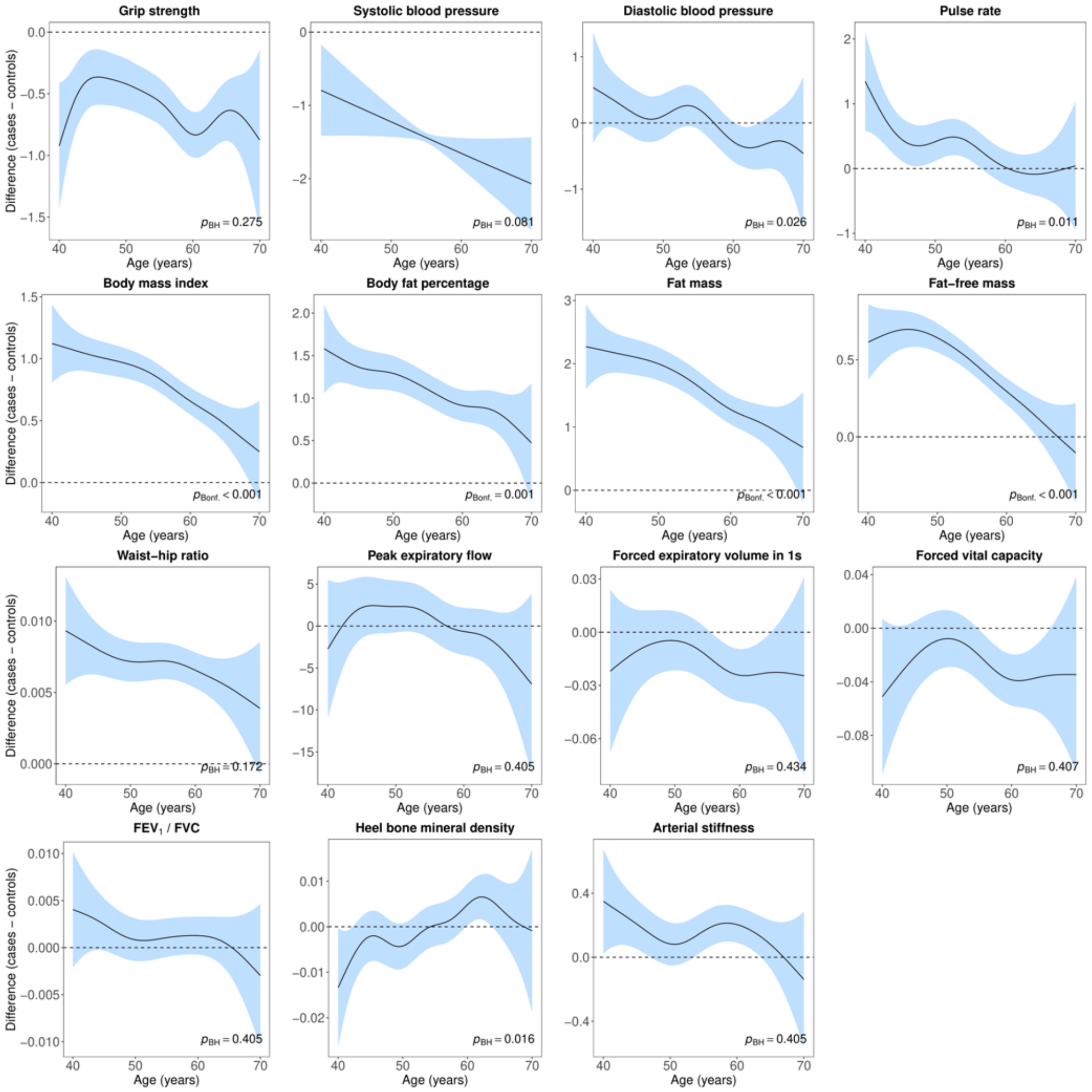

**Supplement figure 7.** Difference smooths comparing age-related changes in physiological measures of females with lifetime depression (**CIDI-SF**) to healthy controls. Models were adjusted for ethnicity (except lung function), gross annual household income, physical activity, smoking status, alcohol intake frequency, sleep duration and, for cardiovascular measures, current use of antihypertensive medications. The shaded areas correspond to approximate 95% confidence intervals (± 2 × standard error). Negative values on the y-axes correspond to lower values in females with lifetime depression compared to healthy controls. The horizontal lines represent no difference between female cases and controls. CIDI-SF = Composite International Diagnostic Interview Short Form; $FEV_1$ = forced expiratory volume in one second; FVC = forced vital capacity; Bonf. = Bonferroni; BH = Benjamini & Hochberg.

8D. Age-related changes in males

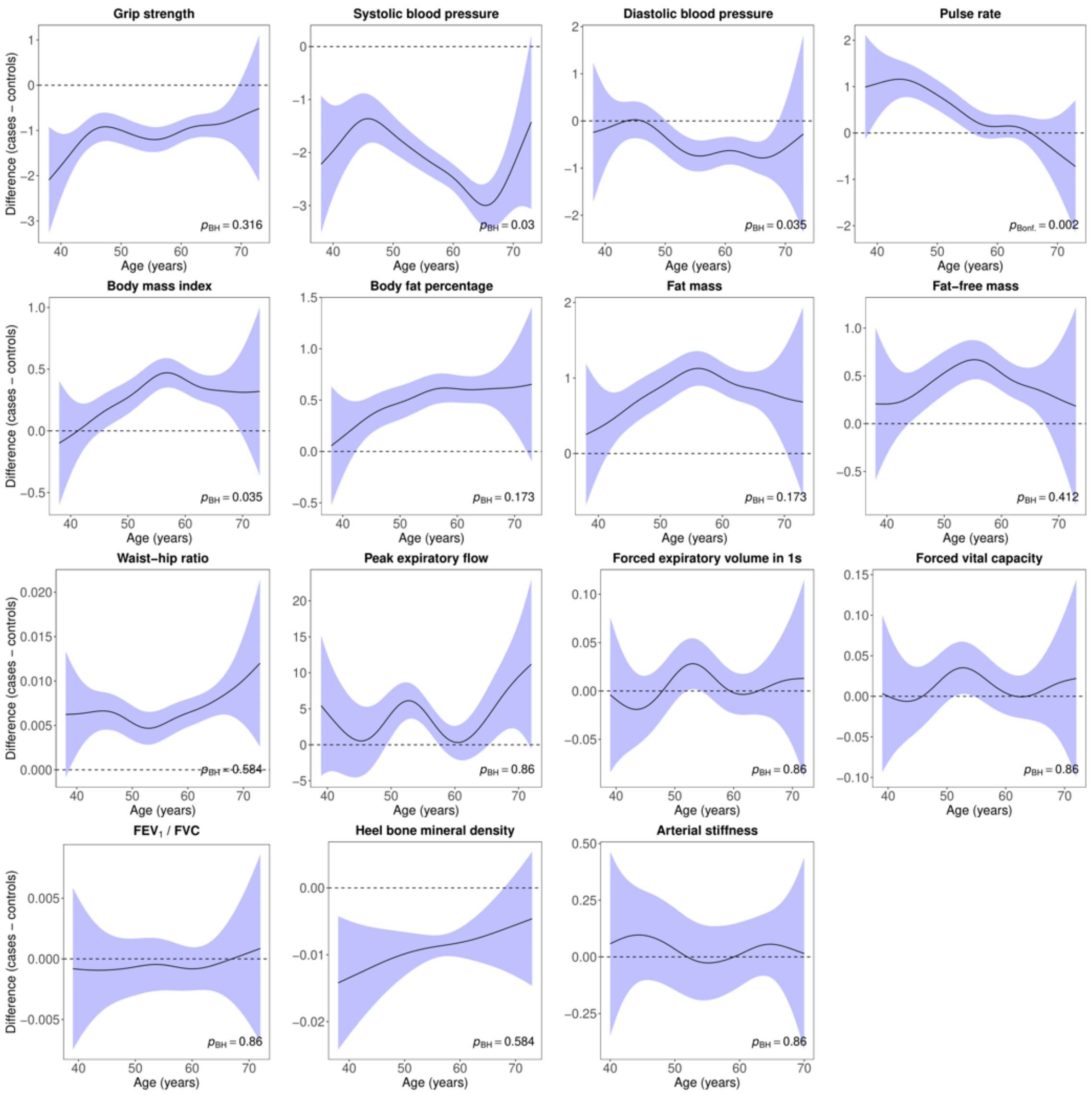

**Supplement figure 8.** Difference smooths comparing age-related changes in physiological measures of males with lifetime depression (**≥ 2 depression measures**) to healthy controls. Models were adjusted for ethnicity (except lung function), gross annual household income, physical activity, smoking status, alcohol intake frequency, sleep duration and, for cardiovascular measures, current use of antihypertensive medications. The shaded areas correspond to approximate 95% confidence intervals (± 2 × standard error). Negative values on the y-axes correspond to lower values in males with lifetime depression compared to healthy controls. The horizontal lines represent no difference between male cases and controls. $FEV_1$ = forced expiratory volume in one second; FVC = forced vital capacity; Bonf. = Bonferroni; BH = Benjamini & Hochberg.

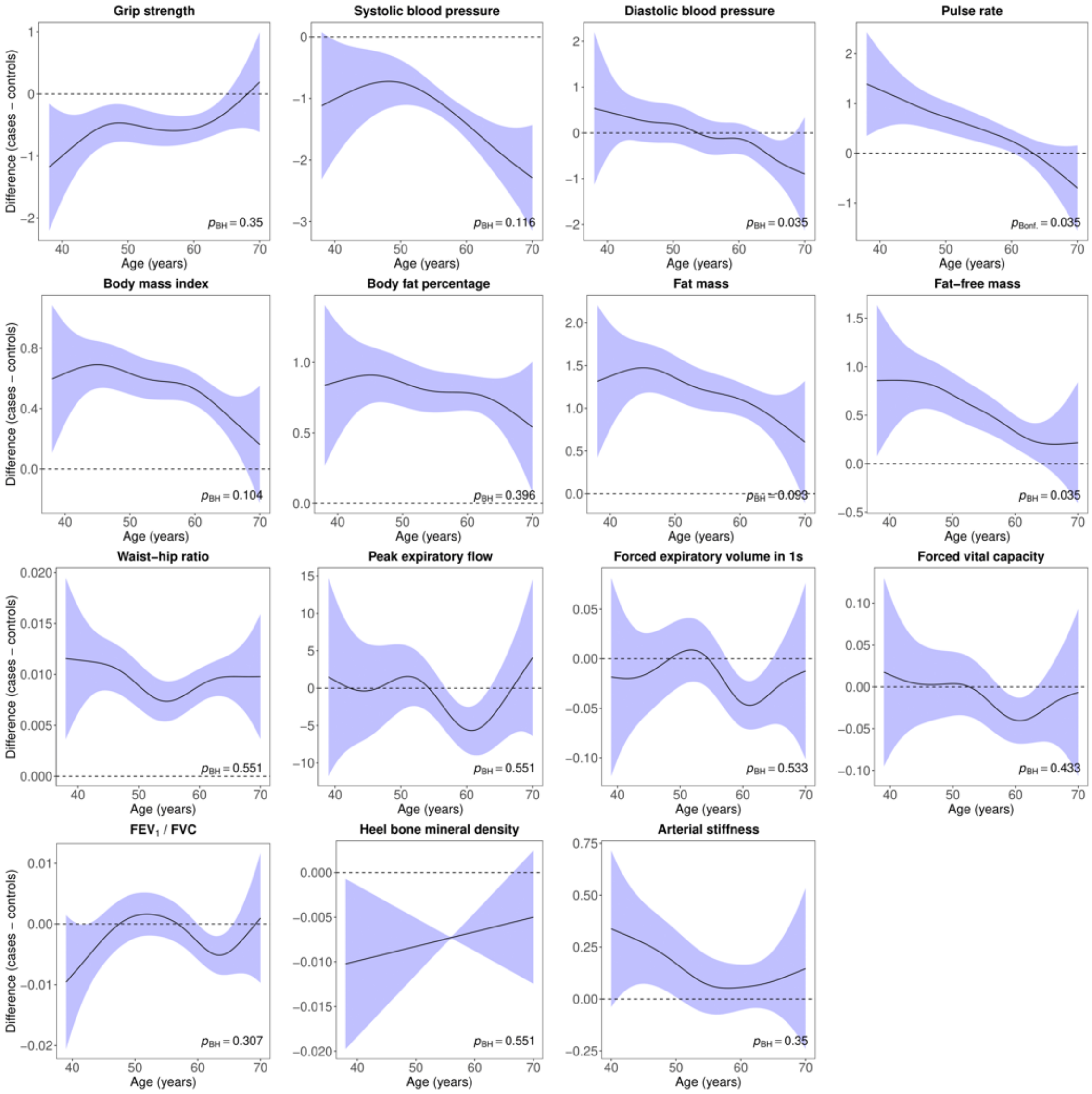

**Supplement figure 9.** Difference smooths comparing age-related changes in physiological measures of males with lifetime depression (**CIDI-SF**) to healthy controls. Models were adjusted for ethnicity (except lung function), gross annual household income, physical activity, smoking status, alcohol intake frequency, sleep duration and, for cardiovascular measures, current use of antihypertensive medications. The shaded areas correspond to approximate 95% confidence intervals (± 2 × standard error). Negative values on the y-axes correspond to lower values in males with lifetime depression compared to healthy controls. The horizontal lines represent no difference between male cases and controls. CIDI-SF = Composite International Diagnostic Interview Short Form; $FEV_1$ = forced expiratory volume in one second; FVC = forced vital capacity; Bonf. = Bonferroni; BH = Benjamini & Hochberg.
